# Genomic surveillance reveals dynamic shifts in the connectivity of COVID-19 epidemics

**DOI:** 10.1101/2023.03.14.23287217

**Authors:** Nathaniel L. Matteson, Gabriel W. Hassler, Ezra Kurzban, Madison A. Schwab, Sarah A. Perkins, Karthik Gangavarapu, Joshua I. Levy, Edyth Parker, David Pride, Abbas Hakim, Peter De Hoff, Willi Cheung, Anelizze Castro-Martinez, Andrea Rivera, Anthony Veder, Ariana Rivera, Cassandra Wauer, Jacqueline Holmes, Jedediah Wilson, Shayla N. Ngo, Ashley Plascencia, Elijah S. Lawrence, Elizabeth W. Smoot, Emily R. Eisner, Rebecca Tsai, Marisol Chacón, Nathan A. Baer, Phoebe Seaver, Rodolfo A. Salido, Stefan Aigner, Toan T. Ngo, Tom Barber, Tyler Ostrander, Rebecca Fielding-Miller, Elizabeth H. Simmons, Oscar E. Zazueta, Idanya Serafin-Higuera, Manuel Sanchez-Alavez, Jose L. Moreno-Camacho, Abraham García-Gil, Ashleigh R. Murphy Schafer, Eric McDonald, Jeremy Corrigan, John D. Malone, Sarah Stous, Seema Shah, Niema Moshiri, Alana Weiss, Catelyn Anderson, Christine M. Aceves, Emily G. Spencer, Emory C. Hufbauer, Justin J. Lee, Karthik S. Ramesh, Kelly N. Nguyen, Kieran Saucedo, Refugio Robles-Sikisaka, Kathleen M. Fisch, Steven L. Gonias, Amanda Birmingham, Daniel McDonald, Smruthi Karthikeyan, Natasha K. Martin, Robert T. Schooley, Agustin J. Negrete, Horacio J. Reyna, Jose R. Chavez, Maria L. Garcia, Jose M. Cornejo-Bravo, David Becker, Magnus Isaksson, Nicole L. Washington, William Lee, Richard S. Garfein, Marco A. Luna-Ruiz Esparza, Jonathan Alcántar-Fernández, Benjamin Henson, Kristen Jepsen, Beatriz Olivares-Flores, Gisela Barrera-Badillo, Irma Lopez-Martínez, José E. Ramírez-González, Rita Flores-León, Stephen F. Kingsmore, Alison Sanders, Allorah Pradenas, Benjamin White, Gary Matthews, Matt Hale, Ronald W. McLawhon, Sharon L. Reed, Terri Winbush, Ian H. McHardy, Russel A. Fielding, Laura Nicholson, Michael M. Quigley, Aaron Harding, Art Mendoza, Omid Bakhtar, Sara H. Browne, Jocelyn Olivas Flores, Diana G. Rincon Rodríguez, Martin Gonzalez Ibarra, Luis C. Robles Ibarra, Betsy J. Arellano Vera, Jonathan Gonzalez Garcia, Alicia Harvey-Vera, Rob Knight, Louise C. Laurent, Gene W. Yeo, Joel O. Wertheim, Xiang Ji, Michael Worobey, Marc A. Suchard, Kristian G. Andersen, Abraham Campos-Romero, Shirlee Wohl, Mark Zeller

## Abstract

The maturation of genomic surveillance in the past decade has enabled tracking of the emergence and spread of epidemics at an unprecedented level. During the COVID-19 pandemic, for example, genomic data revealed that local epidemics varied considerably in the frequency of SARS-CoV-2 lineage importation and persistence, likely due to a combination of COVID-19 restrictions and changing connectivity. Here, we show that local COVID-19 epidemics are driven by regional transmission, including across international boundaries, but can become increasingly connected to distant locations following the relaxation of public health interventions. By integrating genomic, mobility, and epidemiological data, we find abundant transmission occurring between both adjacent and distant locations, supported by dynamic mobility patterns. We find that changing connectivity significantly influences local COVID-19 incidence. Our findings demonstrate a complex meaning of ‘local’ when investigating connected epidemics and emphasize the importance of collaborative interventions for pandemic prevention and mitigation.

## Introduction

Human contact networks can help elucidate SARS-CoV-2 transmission dynamics. For example, it has been shown that the risk of infection for an individual increases with the number of contacts they have[1], the locations they visit[2], and the length of their visit[3], and that the interactions of individuals between different locations hinders virus containment efforts[4–7]. As a result, we would expect SARS-CoV-2 transmission to be higher between geographic locations that have higher human connectivity[8,9]. Reconstructing the interaction networks and temporal dynamics between these “high connectivity” locations can illuminate the sources of emerging waves and help inform intervention strategies and at-risk populations[10]. Genomic data provides one way to measure these connectivity networks, as the geographic spread of rapidly evolving viruses like SARS-CoV-2 can be inferred from molecular data[11,12].

Genomic surveillance programs, many of which were established during the COVID-19 pandemic, have generated large amounts of SARS-CoV-2 genomic data that has been used to track the spread and evolution of the virus in near real-time[13]. While it is clear from genomic data that SARS-CoV-2 spreads between locations causing the reach of local outbreaks to overlap[14–18], we know little about how to quantify this interaction, or the factors contributing to spread. In addition, it is unclear if the temporal dynamic of viral spread has changed as the pandemic transitions from the introductory and expansion phase into endemicity.

The initial spread of SARS-CoV-2 caused the implementation of social distancing policies and international travel restrictions. The former have been studied extensively, showing marked changes in individual behaviors and movements[19,20]. International travel restrictions, enacted by over 89 countries during the first five months of the pandemic[21], stand in contrast to recommendations by the World Health Organization, which argued against the use of travel restrictions and border closures due to their substantial economic, social, and ethical effects, and a lack of evidence on their effectiveness[22]. Although some travel restrictions—including complete border closures, quarantines, and testing requirements—have since been shown to reduce imported cases[17,23–26], the relative contribution of imported cases on local cases before, during, and after restrictions is not fully understood. Additionally, connectivity across heavily-traveled and economically important land-borders, like the US-Mexico border, which was crossed 400 million times a year prior to the pandemic[27] and was closed to non-essential travel from March 19^th^, 2020–November 8^th^, 2021 [28,29], remains unstudied.

In this study, we characterized the connectivity of local COVID-19 epidemics over space and time, how travel restrictions affect this connectivity, and the impact of virus imports on epidemic growth. To reconstruct the dynamics of virus transmission from the beginning of the pandemic to the end of the first Omicron wave (March 2020–December 2022), we sequenced more than 82,000 SARS-CoV-2 samples as part of our routine genomic surveillance in San Diego, California and Baja California, Mexico, and compared them to SARS-CoV-2 genomes from North America and the rest of the world. By studying locations along international borders, we describe the transmission of SARS-CoV-2 across heavily traveled land borders during and after travel restrictions. We find that the implementation and relaxation of travel restrictions resulted in a dynamic shift in transmission of SARS-CoV-2 over the course of the pandemic. Infections resulting from travel accounted for slightly less than half of all COVID-19 cases during epidemic peaks, but up to nearly 80% when cases were lower or during holiday and vacation periods. Our findings indicate that connectivity between locations plays an increasing role in maintaining local epidemics, which highlights the need for collaboration between regional and international governments to enact effective prevention strategies.

## Results

### North American locations experienced similar changes in connectivity over time

To understand how temporal and geographic changes in the connectivity of North American locations influenced SARS-CoV-2 transmission patterns, we investigated SARS-CoV-2 genomic data during the first five waves of the pandemic, until December 2022. We found that locations in North America became more connected over time, likely as a result of the easing of COVID-19 mandates.

We used phylogenetic similarity as a proxy for connectivity, since an increase in connectivity between locations would lead to an increased similarity of their viral populations due to frequent transmission between the locations[30]. Therefore, we measured phylogenetic similarity in the viral populations of epidemics in North American counties, states, or provinces, for each month of the pandemic (**Figure 1A**). To do this, we used the PhyloSor similarity[31] metric, which quantifies the similarity of viral populations as the proportion of branch lengths in a phylogenetic tree that are shared relative to the total branch lengths of both populations[31] (**Supp. Figure 1**). We found that in simulations, PhyloSor similarity recapitulated the number of contacts between communities (see methods; **Supp. Figure 2**) and was thus an appropriate metric to investigate connectivity.

**Figure 1.**
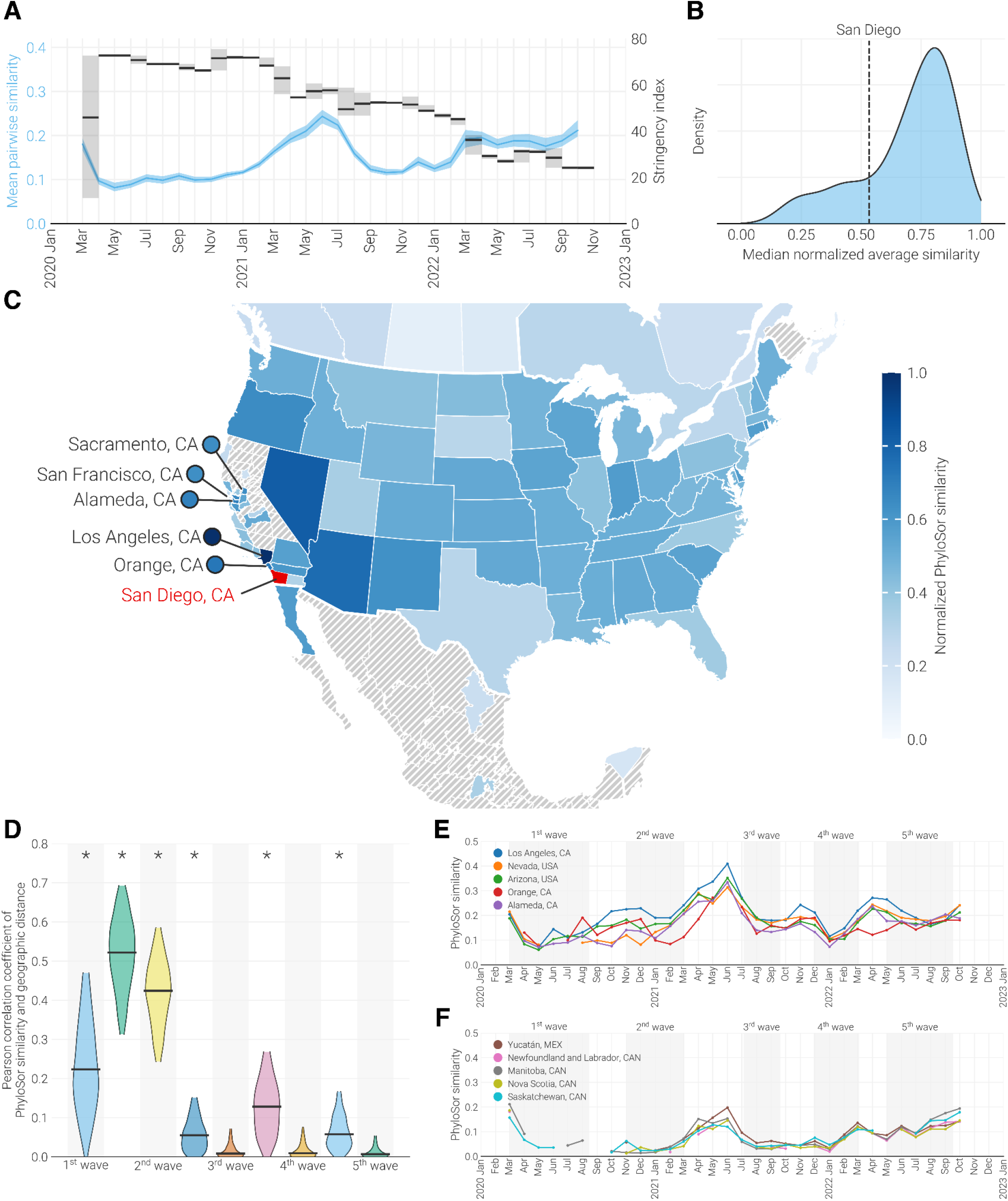
Regional similarity to SARS-CoV-2 genomes collected in San Diego. (**A**) Primary axis, (blue, indicates temporal trends in the mean pairwise PhyloSor similarity of North American locations. Shaded region indicates 95% confidence interval as calculated by bootstrapping locations 100 times. Secondary axis, in black shows the mean stringency of the US government’s response to COVID-19. Higher values indicate a stricter response. Shaded area refers to the range of stringency values observed in a given month. (**B**) Distribution of median min-max normalized average PhyloSor similarity for all locations in North America. The median normalized phylogenetic similarity of San Diego to all other locations is indicated by the dashed vertical line. (**C**) Map showing each location’s median min-max normalized PhyloSor similarity to San Diego for the period of March 2020–August 2022. Here location refers to the county level within California and the state level in the rest of the United States, Canada, and Mexico. Each location is colored by their median value, and locations which were not included in the analysis are hashed out in gray. San Diego is indicated in red. Some parts of Canada, Mexico, and the United States are excluded for clarity. (**D**) Pearson correlation coefficient between median PhyloSor similarity to San Diego and log-normalized centroid-centroid distance to San Diego for each period of the pandemic. Waves of cases are indicated by a gray box, while troughs reside between successive waves. Wave definitions can be found in **Supp.** Figure 6. Confidence intervals calculated by bootstrapping 1000 times. An asterisk indicates that the p-value of correlation is less than 0.05. (**E-F**) Temporal differences in PhyloSor similarity to San Diego for the 5 locations with the highest (D) and lowest (E) median normalized PhyloSor similarity to San Diego.

Prior studies have found that local COVID-19 epidemics were regionally connected during the beginning of the pandemic[15–17], when travel restrictions and social-distancing measures were the most stringent. Our PhyloSor analysis showed that North America had highly similar virus populations at the onset of the pandemic, but we found that virus populations grew increasingly divergent, until they started to become more similar again after May 2020 (**Figure 1A** & **Supp. Figure 3**). This trend was negatively correlated with the stringency of COVID-19 mandates as measured by the Oxford COVID-19 Government Response Tracker[32], suggesting that the relaxation of restrictions was associated with increased connectivity of North American locations (Pearson r = -0.68 [95% CI: -0.43 to -0.83]; P < 0.001).

To better understand factors that explain this trend, we performed detailed analyses of SARS-CoV-2 diversity at a local scale, where transmission and mobility patterns could be more easily interpreted. We used results from our PhyloSor analysis to rank North American counties, states, and provinces by their average phylogenetic similarity to all other locations over time. We found that the average phylogenetic similarity of San Diego County (henceforth San Diego) did not fall in either extreme (18^th^ percentile) and was representative of the widespread trend in connectivity (**Figure 1B**).

### Connectivity of local COVID-19 epidemics became more widespread over time

In addition to being representative of US connectivity trends, San Diego is a popular vacation destination[33] and, along with Tijuana, Baja California, Mexico, contains the busiest international border crossing in the Western Hemisphere[34]. To investigate widespread trends in the diversity of SARS-CoV-2 at the local level, we generated and analyzed 80,323 and 1,950 SARS-CoV-2 genomes from San Diego and Baja California, respectively, making San Diego one of the most densely sampled locations in the United States (collection dates from March 25^th^, 2020 to December 13^th^, 2022; **Supp. Figure 4**).

To investigate temporal dynamics of connectivity before, during, and after the implementation of COVID-19 mandates, we quantified the phylogenetic similarity of SARS-CoV-2 from San Diego with other counties in California and North American states throughout the pandemic. We found that the phylogenetic similarity of SARS-CoV-2 genomes from San Diego were closer to nearby locations, such as Los Angeles, Orange, and Alameda counties, as well as Arizona and Nevada (**Figure 1C**). However, we found only a weak correlation between each location’s median phylogenetic similarity to San Diego and its geographic distance to San Diego (**Supp. Figure 5**; Pearson p value = 0.003; r^2^ value = 0.10). The high similarity to neighboring locations despite the lack of a strong correlation between geographic distance and phylogenetic similarity indicates that locations were only consistently connected to neighboring locations throughout the pandemic.

We hypothesized that the lack of a strong correlation between geographic distance and phylogenetic similarity resulted from an increase in the long range connectivity of locations following the relaxation of COVID-19 mandates. To test this, we calculated the correlation between phylogenetic similarity and geographic distance to San Diego for all locations through each of the five waves of the COVID-19 pandemic (**Supp. Figure 6** & **Figure 1D**). We found that phylogenetic similarity was correlated with geographic distance during the first two waves, but not during the following three waves (Pearson p value < 0.05 for 1^st^-2^nd^ waves; r^2^ > 0.15 for 1^st^-2^nd^ waves; **Figure 1D**). This shows that the connectivity of locations increased in geographic reach after the first two waves of the pandemic.

In contrast to the waves of the pandemic, we noticed that a significant correlation between phylogenetic similarity and geographic distance persisted during the periods of low COVID-19 incidence (**Figure 1D**). This led us to investigate the association between connectivity and COVID-19 incidence. We found that, even after subsampling to correct for potential sampling biases (**Supp. Figure 7**), similarity between San Diego and other locations increased during periods of low COVID-19 incidence relative to the adjacent waves (**Figure 1E-F**). The effect was most pronounced in the locations with the highest phylogenetic similarity to San Diego, suggesting that increased viral diversity during periods of high incidence was responsible for the observed reduction in similarity between locations.

### SARS-CoV-2 population similarity is driven by transmission frequency

A key component of understanding transmission dynamics is determining if viral similarity between locations is due to transmission between them (“bidirectional transmission”) or shared introduction sources. To understand the factors that drove phylogenetic similarity, we estimated SARS-COV-2 transmission between locations by reconstructing the timing and number of geographic transitions (also called Markov jumps[35]) into and out of San Diego across the full posterior of a Bayesian phylogeographic reconstruction (**Figure 2A**). Despite all locations having similar numbers of genomes in the phylogeny (see Methods), we observed the largest number of transitions between San Diego and Los Angeles (**Figure 2B**), indicating that bidirectional transmission frequency was consistent with viral similarity.

**Figure 2.**
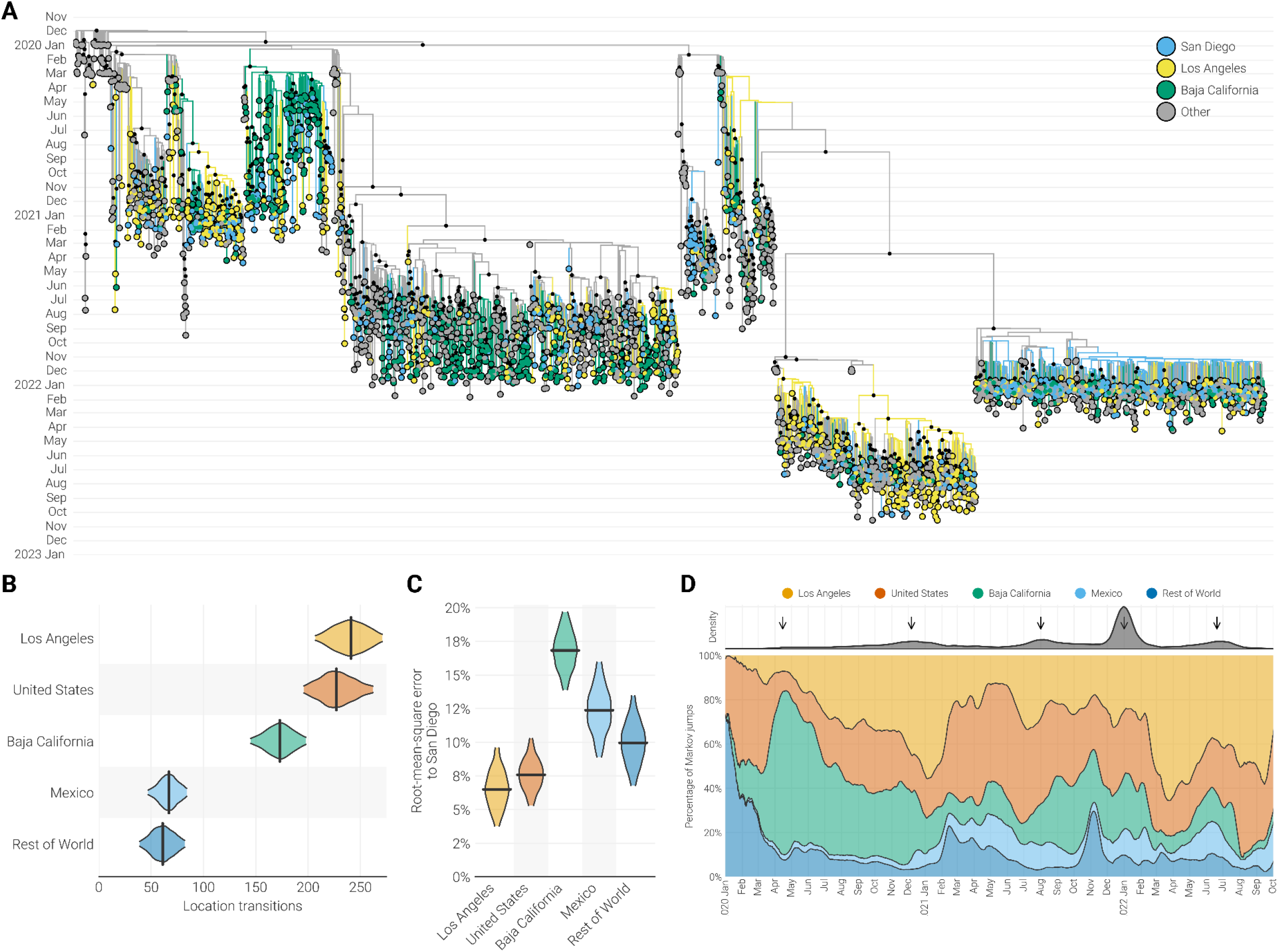
Phylogenetic analysis of SARS-CoV-2 in the Californias. (**A**) Maximum clade credibility tree of whole genome SARS-CoV-2 sequences sampled from Baja California, Los Angeles County, San Diego, and the rest of the world. Black circles at internal nodes indicate posterior support greater than 0.5. (**B**) Median number of transitions between each location and San Diego inferred by phylogeographic reconstruction. Black bar indicates the median value. (**C**) Root-mean-square error between the estimated source composition of introductions into each location compared to San Diego. (**D**) Proportion of location transitions between San Diego and all other locations in the discrete state analysis. Top facet indicates the temporal density of location transitions across the posterior distribution of trees. Arrows are used to show periods of increased location transitions.

To determine if shared introduction sources also contributed to similarity between San Diego and locations highly connected to it, we calculated the percentage of transmission into each location that originated from each other location across the posterior distribution of trees (i.e. the introduction profile; **Supp. Figure 8**). We found that Los Angeles and San Diego had more similar introduction profiles than Baja California and San Diego (Los Angeles vs. San Diego introduction profile root-mean-square error [RMSE]: 6.5 percentage points [95% HPD 3.8-9.6 points]; Baja California vs. San Diego introduction profile RMSE: 16.8 points [95% HPD 13.9-19.8 points]; **Figure 2C**). This suggests that bidirectional transmission played a larger role than shared introduction sources in driving similarity between San Diego and Baja California, though both contributed to the similarity between San Diego and Los Angeles. As a result, bidirectional transmission is a better representative of the connectivity between locations than phylogenetic similarity.

We next examined the temporal dynamics of bidirectional transmissions between San Diego and other locations. Using the same phylogeographic reconstruction, we found that bidirectional transmission involving San Diego was consistently present over time, but increased during five periods: (1) April–May 2020, (2) November–December 2020, (3) July–September 2021, (4) December 2021–February 2022, and (5) June–July 2022 (**Figure 2D**). The earliest period was dominated by transmission with Baja California, whereas during later periods transmission across the border was largely replaced by transmission with Los Angeles and other farther domestic locations (**Figure 2D**). This result is in agreement with findings from our PhyloSor analysis, suggesting that the frequency of transmission between distinct locations increased during the pandemic.

To examine how bidirectional transmission impacted local COVID-19 incidence, we investigated the temporal association of connectivity and local case numbers. We estimated the relative amount of incidence that could be attributed to connectivity as the percentage of viral lineages circulating in San Diego that could not be traced back to lineages circulating in San Diego at least two weeks earlier in our phylogeographic reconstruction. We found that, on average, half of all lineages in San Diego could be attributed to other locations, but the proportion decreased markedly during the waves of the epidemic relative to the adjacent troughs (**Supp. Figure 9**). This result is supported by contact tracing data, which indicated that, on average, 15% of all San Diego cases were directly associated with travel within the US or Mexico, including from Los Angeles and Baja California, and that the percentage of cases directly associated with travel was inversely correlated to COVID-19 incidence (**Supp. Figure 10**; Pearson r = -0.23 [95% CI: -0.03 to -0.42]; P = 0.03).

Combined, these results indicate that connectivity between locations played a prominent role in maintaining local incidence, particularly during periods between epidemic waves, sustaining the COVID-19 pandemic.

### Temporal shifts in mobility impacted SARS-CoV-2 transmission risk

Our observation that connectivity to nearby locations impacted local COVID-19 cases prompted us to investigate factors driving transmission. Investigations of prior outbreaks have shown that a gravity model[14,15,17], wherein the spread of a pathogen between locations is a function of the mobility between them and the infection rate at the origin, can be used to estimate transmission. We asked if the spread of SARS-CoV-2 could also be explained by this model, and ultimately found that mobility was an important driver of SARS-CoV-2 transmission and that changes in mobility corresponded to the increases in connectivity observed in previous analyses.

To see if the gravity model explains SARS-CoV-2 spread, we examined whether transmission between locations with high connectivity followed mobility patterns estimated by travel surveys[36] and was correlated with the estimated number of infections at the origin. We found that SARS-CoV-2 lineages transmitted to San Diego from Baja California were more likely to be introduced in the San Diego regions closest to the border (**Figure 3A-C**). This aligned with travel surveys conducted within San Diego prior to the pandemic that reported that visitors from Mexico traveled on average less than 30 miles in the US and typically remained within the border regions of San Diego[36]. Additionally, we found that transmission between San Diego and Los Angeles, and between San Diego and Baja California, was correlated with the estimated number of asymptomatic infections in that location that were able to travel from the source location (only infections that are pre-symptomatic or asymptomatic, see Methods; r^2^ > 0.59 for all pairs; **Figure 3D-G**). Our findings that there is a clear correlation between transmission magnitude and infection rate, together with the concordance between transmission across the US-Mexico border and known mobility patterns, indicate that the gravity model accurately describes SARS-CoV-2 transmission.

**Figure 3.**
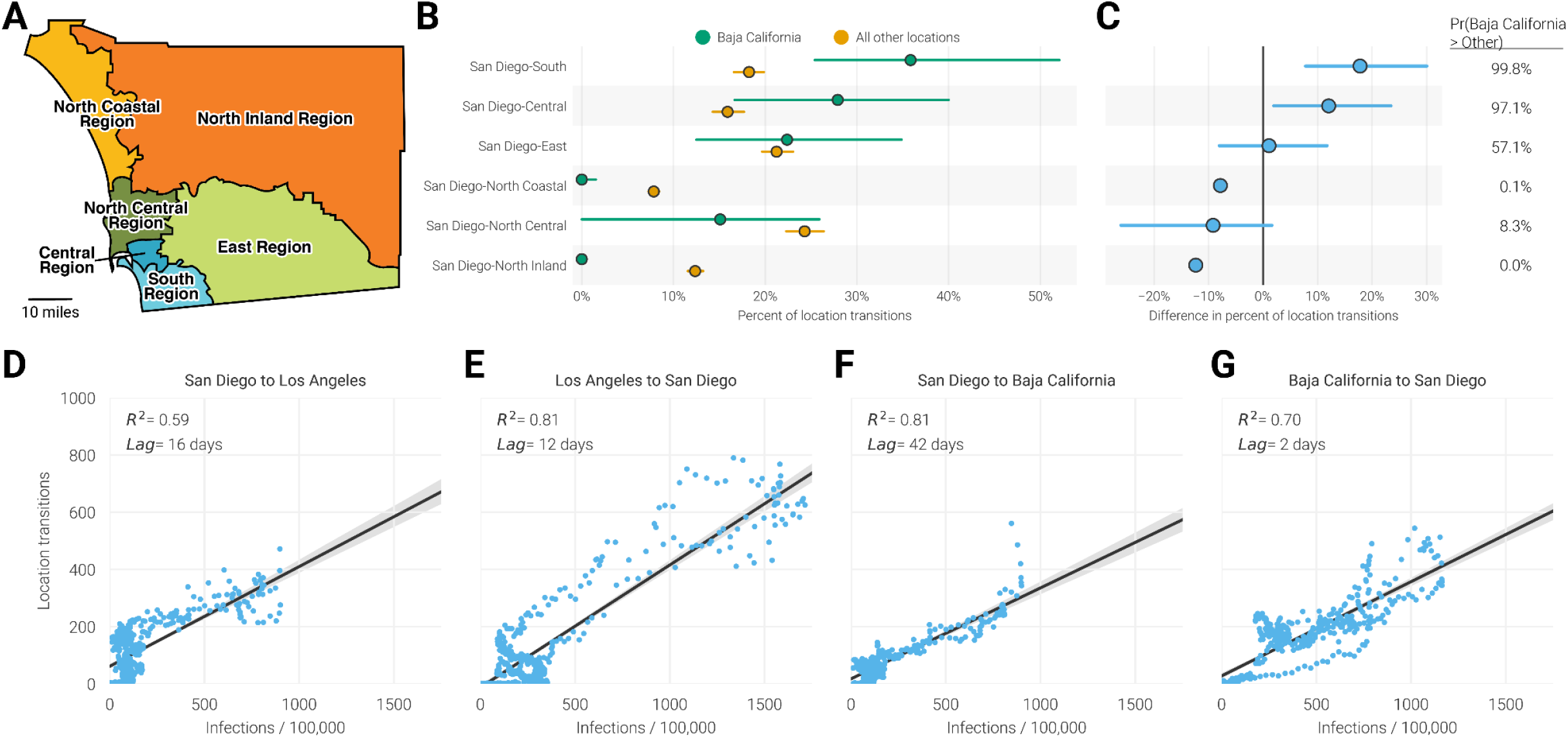
Dynamics of cross-border transmission. (**A**) Boundary of approximate Health and Human Service Agency (HHSA) regions within San Diego. Scale bar indicates a distance of 10 miles. (**B**) Percentage of location transitions from either Baja California (in green) or all other locations (in orange) into San Diego that were inferred to land in each of the county’s HHSA regions. Dots indicate the median value while bars show the 95% highest posterior density interval. (**C**) Relative difference in percentage of location transitions originating in either Baja California or outside Baja California for each of San Diego’s HHSA region indicated in panel **B**. Probability refers to the percentage of trees in the posterior in which the proportion of location transitions from Baja California is greater than the proportion from all other locations combined. (**D-G**) Correlation between the magnitude of location transitions and the estimated number of infections at the origin for each location pair indicated. R^2^ was determined using ordinary least squares regression. In order to limit the impact of vaccines on our infection calculations, we only show the correlation for dates prior to May 5th, 2021 when at least 50% of San Diego’s population received at least one dose of a SARS-CoV-2 vaccine[37,38].

Using the gravity model, we next assessed how changes in mobility over time impacted SARS-CoV-2 transmission. To examine the relationship between mobility and transmission, we analyzed weekly land and air travel data generated by SafeGraph[39]. We found that neighboring California counties (Riverside, Los Angeles, and Orange), states (Arizona), and countries (Mexico; of which 99% originates from Baja California[36]) consistently dominated mobility into San Diego (**Figure 4A**). Further, we found that there was a moderate correlation between the number of travelers arriving from a location and the median phylogenetic similarity of that location to San Diego (Pearson r = 0.44 [95% CI, 0.24 to 0.60]; P < 0.001; **Figure 4A** & **Supp. Figure 11**). Consistent with our previous findings that connectivity to more distant locations increased over the course of the pandemic, we found that mobility from more distant locations also increased over time. (**Figure 4B-C**). Travelers from Riverside County, Los Angeles County, Orange County, Arizona, and Mexico accounted for 75% of travelers into San Diego during early 2020, but less than 50% of travelers from June 2021 onwards (**Figure 4C**). This latter amount is similar to the proportion of travelers arriving from these locations during 2019, before responses to the COVID-19 pandemic affected mobility (**Figure 4C**).

**Figure 4.**
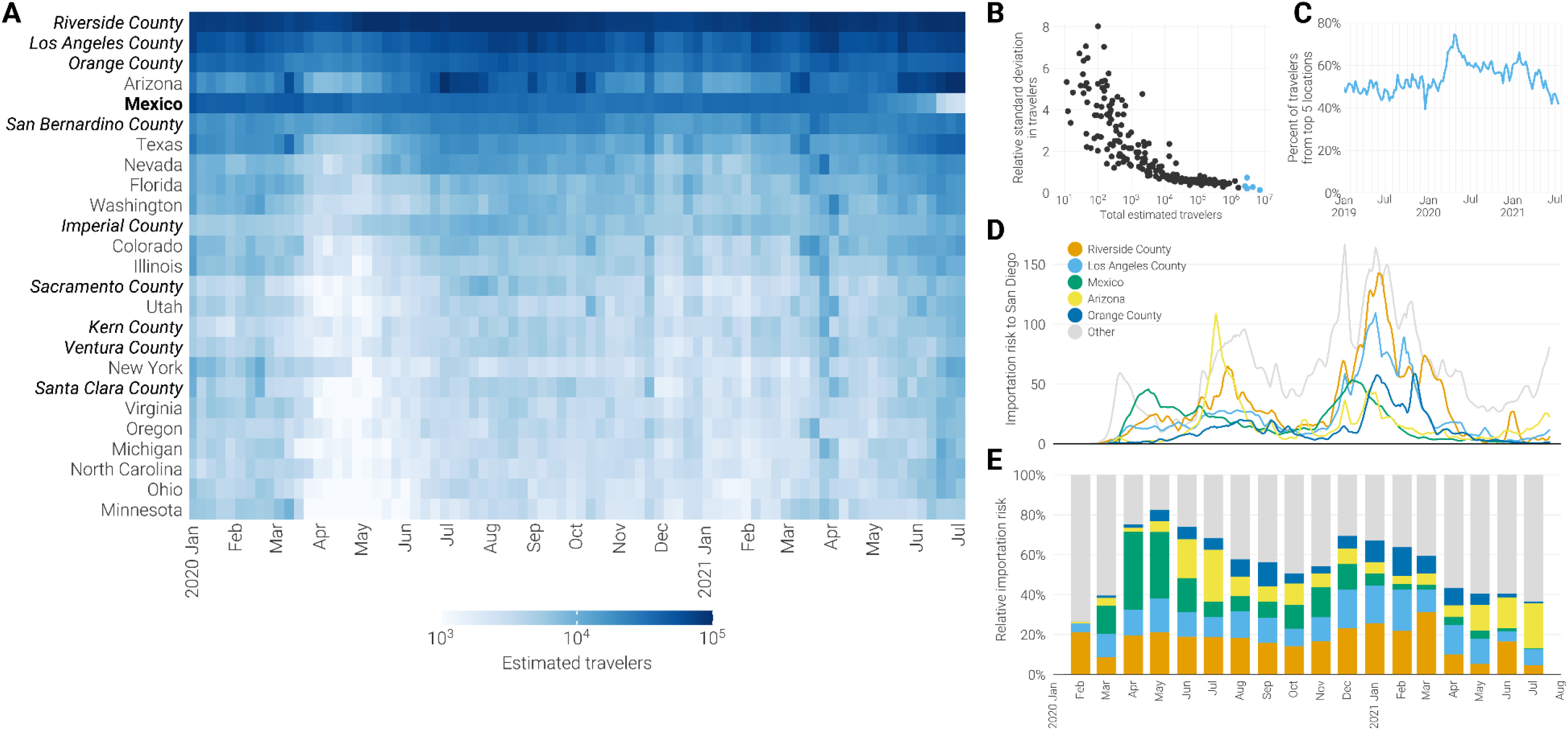
SARS-CoV-2 import risk into San Diego. (**A**) Weekly estimated number of travelers arriving into San Diego from January 2020–June 2021. Locations are sorted by the total number of estimated visitors over this period and only the top 25 are shown. Location names are styled depending on their administrative level: California counties are italicized, countries are bolded, and US states weighted normally (**B**) Scatter plot of each locations’ total estimated travelers into San Diego and the relative standard deviation in estimated travelers for the period indicated by panel **A**. The five locations with the greatest total estimated travelers into San Diego are highlighted in blue. (**C**) Proportion of travelers arriving into San Diego from the five locations with the greatest total estimated travelers (top-most five locations in panel **A**). (**D**) import risk into San Diego. import risk was estimated based on the number of infectious travelers relative to the population size and the total number of travelers at the origin. Only the five locations with the greatest total import risk into San Diego are shown. All other locations are colored in gray. (**E**) Relative import risk into San Diego. Locations are colored as in panel **D**, with gray representing all locations outside the top five locations.

We then considered the infection rate at the transmission source location, the other component of the gravity model[40], by estimating the number of COVID-19 infected travelers arriving into San Diego from each location (“import risk”). We calculated this as the product of the number of travelers arriving in San Diego from each source location as determined by SafeGraph data and the estimated COVID-19 infection rate at the source location (**Figure 4D**). We found that the relative import risk from neighboring locations was low at the beginning of the pandemic, peaked during the spring of 2020, and decreased steadily from then until the end of available data (74.8% in April 2020 to 36.9% in July 2021; **Figure 4E**). This trend parallels changes in mobility and transmission estimates (**Figure 2D** & **Figure 4C**), supporting our findings that the connectivity and major sources of imports into local epidemics shifted from neighboring locations to more distant domestic locations as a result of the relaxation of COVID-19 mandates, such as stay-at-home orders, curfews, and business closures.

### US-Mexico border closure was ineffective in preventing imported cases

Our analyses indicate that there was significant transmission between San Diego and nearby locations, including Baja California, during the entire duration of the pandemic. However, the US-Mexico border was closed to non-essential travel from March 19^th^, 2020–November 8^th^, 2021[28,29], prompting us to evaluate whether this restriction was effective at preventing cross-border transmission. To do so, we measured how changes in mobility resulting from the border closure impacted import risk into San Diego.

Using mobility data from SafeGraph, we found that the number of northbound travelers across the US-Mexico border was 23.1% less during the partial closure (March 2020 to July 2021) than in 2019 (**Supp. Figure 12**). We additionally found that import risk across the border was reduced by 22.8% when we compared the observed import risk from Mexico to import risk calculated using mobility estimates from 2019 (**Supp. Figure 12**). However, given that the majority of travelers into San Diego arrived from locations other than Mexico (**Figure 4A**), this reduction only amounted to a 3.1% reduction in the total import risk into San Diego, indicating the impact of the non-essential closure was ineffective (**Figure 5A**). However, we observed a general reduction in the number of travelers visiting San Diego from most locations beginning in March 2020, which were affected by stay-at-home orders and travel hesitancy rather than official travel restrictions[3,24] (**Figure 4A**). This prompted us to study whether the reduction in mobility from Mexico to San Diego was more or less impactful than general reductions in mobility.

**Figure 5.**
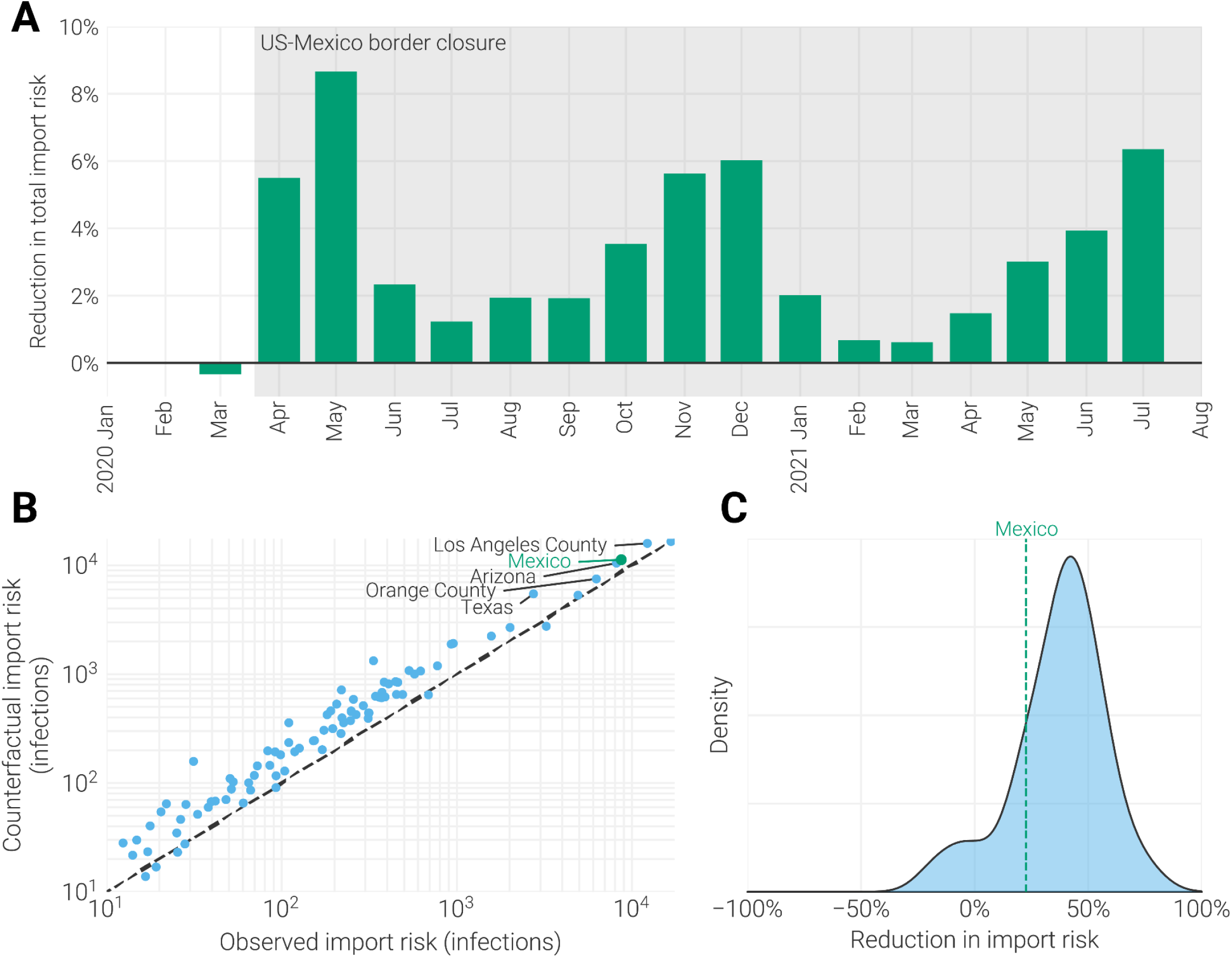
Impact of US-Mexico border closure. (**A**) Percentage increases in the total import risk into San Diego when travel from Mexico is held at 2019-levels compared to observed travel. (**B**) Plot of total counterfactual import risk (calculated using mobility estimates from 2019) vs. observed import risk for all locations with an absolute import risk greater than 10 infected travelers (accounting for 45% of all locations and 99.8% of the total import risk into San Diego). The five locations with the greatest difference between the counterfactual and observed import risk are labeled. import risk from Mexico is indicated with a green point. (**C**) Distribution of the relative reduction from the counterfactual to the observed import risk for each location in panel **B**. Mexico’s relative reduction of 22.8% is indicated by the dashed vertical bar.

To investigate this, for each location, we compared the observed import risk to San Diego with import risk calculated using mobility estimates from 2019 (**Figure 5B**). While we found that Mexico had the 3^rd^ largest reduction in import risk, behind Los Angeles County and Texas, the reduction was only a small fraction of the total import risk from Mexico (22.8% reduction). Specifically, we found that the relative reduction in import risk from Mexico was less than 80.0% of all other locations and half of the median reduction in import risk (40.1%; **Figure 5C**). This indicates that the closure of the US-Mexico border to non-essential travel was significantly less impactful at reducing imports than the reductions in travel associated with stay-at-home orders and decreases in mobility resulting from hesitancy[3,24].

## Discussion

In this study, we determined a widespread shift in the connectivity of local COVID-19 epidemics during the pandemic, by integrating genomic surveillance data with epidemiological and mobility data. Focusing on the first five waves of the pandemic, we found that the implementation of COVID-19 mandates, such as travel restrictions and stay-at-home orders, contained the spread of SARS-CoV-2 locally at the beginning of the pandemic. However, the lifting of mandates enabled the virus to spread further as travel increased. We found that travel-associated infections accounted for half of the incidence during some periods of the pandemic, indicating that local outbreaks were largely affected by epidemics in other locations. By estimating cross-border transmission of SARS-CoV-2, we show that closures to non-essential travel were ineffective at preventing transmission, especially when compared to less-targeted restrictions.

We focused on the epidemic in and around San Diego and the US-Mexico border. This region was selected due to its propensity for domestic and international travel, as well as our genomic surveillance in the area. While caution should be taken in applying our conclusions to less-populated or less-connected regions, our PhyloSor analysis suggests that North American counties, states, and provinces similarly experienced the observed shift in connectivity (**Figure 1B**). Other locations and continents underwent distinct patterns of restriction implementation and relaxation[41–43], and thus further work is necessary to determine how these patterns differentially impacted connectivity.

Our finding that local COVID-19 epidemics are highly interconnected highlight the importance of collaborative and inclusive public health measures. The beginning of the pandemic led to a substantial reduction in long distance travel in favor of local connections (**Figure 4A**). This phenomenon was also observed in the national mobility networks of France, Italy, and the UK following their respective lockdowns[44]. The local connections of San Diego extended to neighboring epidemics throughout the pandemic, supporting previous evidence that connectivity between adjacent locations precludes virus containment. For instance, the dispersal of SARS-CoV-2 from Wuhan progressed mainly to adjacent cities[45], and B.1.1.7 was seen to spread from Kent and Greater London to other locations in the UK at a rate proportional to their mobility with Kent and Greater London[46]. San Diego’s connectivity to Mexico, provides further evidence that state and international borders did not act as barriers to the spread of the virus, and indicates that both domestic and international collaborations are necessary to control the spread of pathogens.

When we evaluated enacted control measures, we found that the closure of the US-Mexico border was ineffective at reducing cross-border transmission. Preventing transmission requires completely halting travel whereas the border closure only restricted non-essential travel. While our data provides only limited examples of any other official travel restrictions, we found that the US-Mexico border closure (22.8% reduction in import risk) was less effective than the total border closure in Jordan[17] (65% reduction in import risk), the mandatory 14-day quarantine enacted in Hong Kong[25] (94% reduction in imported cases), and the ban in Wuhan on all outgoing travel[24] (74% reduction in exported cases). Additionally, whereas other non-pharmaceutical interventions caused individuals to take shorter, less complex trips and reduce person-to-person contacts, our finding that the border closure did not result in any changes in the destination of northbound travelers (**Figure 3B**), suggests that the closure had a limited effect on behavior[3]. As a result, the enacted border closure would have had to be much more stringent to be as effective as other travel restrictions in reducing imports of the virus. However, our finding that outbreaks are increasingly interconnected adds to a growing body of evidence that targeted travel restrictions have limited practical value[5,47,48].

Our ability to detect cross-border transmission was due to the pooling of resources and collaboration between academic laboratories, public health laboratories, and hospitals on both sides of the border. A recently developed framework for identifying transmission lineages using limited sequencing resources, indicates that only 0.5% of infections need to be sequenced to detect 95% of transmission lineages with a frequency of at least 2% in the population[49]. Current sequencing efforts in Baja California and San Diego, with a sampling fraction of 1% and 10%, respectively, appear to surpass this recommendation. However, the high lag observed between infections in San Diego and transitions to Baja California (**Figure 3F**), suggests that we detected Baja California transmission lineages later than would be expected given our sampling rate. The difference in sampling rates between San Diego and Baja California, as well as the absence of estimates for the number of southbound travelers crossing the border, prevent any conclusion on the directionality of cross-border transmission[50]. Additionally, other well-traveled border crossings along the US-Mexico border fall even shorter of the recommendation to sequence 0.5% of cases, limiting the region’s ability to monitor disease spread. Considering these locations are large sources of potential infections, it is critical that regional surveillance capacity is strengthened in these areas.

As the COVID-19 pandemic progresses to endemicity, durable genomic surveillance systems will be critical to provide insights into the continued spread and evolution of SARS-CoV-2. We show that the information produced by epidemiological and genomic surveillance can be integrated with mobility data to quantifiably provide estimates on the sources of introductions and local transmission, and how they change over time. We found that for well-connected locations such as San Diego, transmission resulting from connectivity blurred the division between seemingly separated local epidemics, particularly when cases were low in any of the locations. Consequently, it is a necessity that the international community equitably distribute surveillance infrastructure and enact travel restrictions collaboratively. It is vital that the effects of such restrictions, particularly when they are not equally experienced, be carefully weighed against their quantitative benefit.

## Methods

### SARS-CoV-2 Amplicon Sequencing

SARS-CoV-2 RNA samples were collected from routine diagnostic tests performed by SEARCH in San Diego, and by Salud Digna, Centro de Diagnóstico COVID-19, Institute of Epidemiological Diagnosis and Reference, Genomica Lab Molecular, and Infectolab in Baja California, Mexico. SARS-CoV-2 was sequenced using PrimalSeq-Nextera XT. This protocol is based on the ARTIC PrimalSeq protocol[51], except that amplicon sizes were reduced to enable 2x150 read length requirements. The ARTIC network nCoV-2019 V4 primer scheme uses two multiplexed primer pools to create overlapping 250 bp amplicon fragments in two PCR reactions. Full details of protocol can be found here:

[Protocol for HCoV-19 sequencing: PrimalSeq-LibAmp](https://docs.google.com/document/d/1zog56u3UkDoeh-OVS0UsK2SNClHX3amEOmubKl4FC_Q/edit). Briefly, SARS-CoV-2 RNA (2 mL) was reverse transcribed with LunaScript RT (New England Biolabs). The virus cDNA was amplified in two multiplexed PCR reactions (one reaction per primer pool; custom primer scheme can be found here:

[Primers for SARS-CoV-2 PrimalSeq-LibAmp](https://docs.google.com/spreadsheets/d/1iNW68ds8G70JxggeiCaMIkPTechPV9PgO4J9Ak-XMus/edit#gid=0)) using Q5 DNA High-fidelity Polymerase (New England Biolabs). Following an AMPureXP bead (Beckman Coulter) purification of the combined PCR products, sequencing adaptors containing sample specific indexes were added using a step-out PCR reaction using Q5 DNA High-fidelity Polymerase. The libraries were purified with AMPureXP beads and quantified using the Qubit High Sensitivity DNA assay kit (Invitrogen) and Tapestation D5000 tape (Agilent). The individual libraries were normalized and pooled in equimolar amounts at 1.5 nM. The 2 nM library pool was sequenced on an Illumina NovaSeq 6000 (300 cycles kit). Consensus sequences were deposited on GISAID and Raw reads were deposited under BioProject accession ID PRJNA612578.

### SARS-CoV-2 Genomic Data

We queried the GISAID EpiCoV database for all SARS-CoV-2 genomes collected up to January 4^th^, 2023[13]. We removed genomes that (1) were less than 20,000 nucleotides in length, (2) had greater than 12.5% ambiguous nucleotides, (3) had an incomplete or incorrect year-month-day sampling date reported, (4) had a sampling country that could not be interpreted, (5) were not collected from a human infection, (6) had less than 50% agreement with Hu-1 (GenBank Accession ID: NC_045512.2), or (7) had greater than 500 discrete indels. The final dataset contained 13,722,590 genomes.

### PhyloSor Analysis

We used the global SARS-CoV-2 phylogeny provided by GISAID as of January 5^th^, 2023 (also called Audacity). The phylogeny contains all SARS-CoV-2 genomes available on GISAID that were marked as both ‘complete’ and ‘high-coverage’ by GISAID, were longer than 28,000 nucleotides, contained less than 1,000 ambiguous nucleotides, were not identified as being on a long branch, were not manually identified as questionable, and were included in the genomic dataset described previously. For each location pair, the phylogeny was pruned to taxa present in the genomic dataset that were collected from either location. Using the pruned tree, for each month in the period of January 2020 to December 2022, the PhyloSor metric was calculated using only sequences collected in that month[31]. Briefly, the PhyloSor metric is calculated as the ratio of branch lengths (in units of per-site substitution rate) that are shared by two sets of tips (*BL_Both_*) compared to the total branch length that is unique to each set of tips (**Supp. Figure 2**).

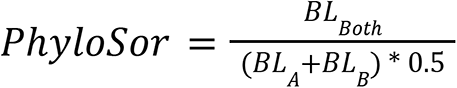

Where BL_A_ and BL_B_ indicate the total branch lengths of either the first set (A) or the second set (B).

To limit the impact of low sampling, we only compared locations that sampled at least 1000 total sequences and collected a sequence in at least 75% of the epidemiological weeks between March 2020 and December 2022. Additionally, within these comparisons we only considered months where at least 30 sequences were included from each location. Here location refers to counties within California, and states in the rest Canada, Mexico, and the US.

To assess differences in PhyloSor similarity resulting from unequal sampling fractions, we compared San Diego’s similarity to all suitably sampled locations under two different subsampling schemes. In the first, a constant number of San Diego sequences were sampled for each month in the analysis, equal to the number of sequences available for San Diego from the month with the least amount of sequences greater than 30. This number was 149. In the second scheme, a number of San Diego sequences were sampled such that they represented a constant fraction of cases. 2.5% was selected as it was the 10^th^ percentile of the sampling fraction of all months. Ten replicates of each subsampling scheme were performed, and the median PhyloSor similarity of San Diego to all other locations was compared between the subsampling schemes and the analysis performed with the non-downsampled dataset.

### PhyloSor Validation

To validate the use of PhyloSor in measuring the temporal connectivity between locations, we conducted epidemic simulations using FAVITES V1.1.35[52]. First we generated static contact networks in FAVITES using a modified Barabási-Albert algorithm[53]. We generated two separate 20,000 member communities using the Barabási-Albert algorithm with a mean value of 8 contacts per day. For each community, we calculated intra-community connectivity as the fraction of all possible contacts that were made. Inter-community edges were sampled by randomly deciding for each pair of nodes in different communities if they should be connected by an edge or not. The probability of connecting two nodes in two communities was calculated as a fraction of the average intra-community connectivity. We called this term inter-community connectivity. Ten contact networks were generated using inter-community connectivity values between 0.5 and 0.001 were simulated.

We then simulated a transmission network over each contact network using a Susceptible-Infected-Recovered model. The simulation sampled a single viral lineage from each infected individual at a random point during their infectious period to represent viral genome sequencing. Each virus phylogeny in units of time (years) was sampled under a constant coalescent model using the Virus Tree Simulator package embedded in FAVITES. All parameters for the transmission network simulation and viral lineage sample were identical to parameters used in Worobey et al. 2020[7]. Ultimately, the virus phylogeny in units of time was converted to units of per-site mutation rate by multiplying the branch length by a constant 1.1 x 10^-3^ subs/site/year, consistent with Duchene et al. 2020[54]. PhyloSor similarity between the two communities for the first month of the simulation was calculated using the phylogeny. We detected a strong correlation between PhyloSor similarity and inter-community connectivity (Pearson r = 0.89 [95% CI: 0.82-0.93]; P < 0.001).

### Network Analysis

For each month in the period of January 2020-November 2022, we considered a complete weighted undirected graph, where nodes are locations in North America and edges weights are the PhyloSor similarity between locations. For this analysis, location refers to the county-level in the US, and state-level in Canada and Mexico. However, where counties did not meet the inclusion criteria (greater than 1000 sequences and at least one sequence in 75% of the epidemiological weeks between March 2020 and December 2022), their sequences were assigned to the state-level. We calculated the average pairwise similarity between all locations as the global efficiency of the graph, which takes into consideration the multiple pathways between locations in the graph. Global efficiency is a network measure that describes how easily information is exchanged over the network and can be defined as the average shortest path length between each pair of nodes in the network[55]. Low global efficiency indicates a network has few strong connections, while high efficiency indicates that most locations are strongly connected. Given a weighted network *G* with *n* nodes, global efficiency can be calculated as:

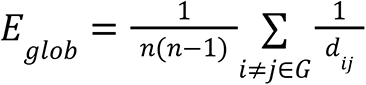

where *d_ij_* is the distance of the shortest path length between nodes *i* and *j*. The shortest path length is the smallest sum of weights throughout all the possible paths in the network from *i* to *j*. In our case, because our edge weights represent a similarity metric rather than a distance metric, we used the reciprocal of edge weights to calculate the shortest path length between nodes.

The contribution of each node to global efficiency, also called nodal efficiency, is the average PhyloSor similarity between it and all other nodes. In our network, locations with high nodal efficiency are phylogenetically similar to a greater portion of North American locations. It can be calculated as:

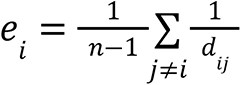

To summarize across the networks of all months, the nodal efficiencies of all locations for a given month were min-max normalized and the median normalized nodal efficiencies were reported.

### Stringency of US Response to COVID-19

To summarize the strictness of the US government’s response to COVID-19, we used the Stringency Index as calculated by the Oxford Coronavirus Government Response Tracker[32]. Briefly, the index is a composite metric which takes into account school closures, workplace closures, cancellation of public events, restrictions on public gatherings, closures of public transport, stay-at-home requirements, public information campaigns, restrictions on internal movements, and international travel controls (see citation for full calculation details). The metric was calculated daily for the US and returns a value between 0 and 100; a higher score indicating a stricter response. We additionally calculated the mean stringency index for each month in the period of January 2020 to November 2022.

### Genomic Dataset Generation

The massive amount of sequencing data produced during the COVID-19 pandemic prevented us from including all data in our phylogenetic analyses. In order to limit the computational burden of the phylogeographic analysis, we subsampled 2500 genomes from our SARS-CoV-2 genomic dataset. To focus the analysis on the region around San Diego County, we allocated 500 genomes each to San Diego County, Los Angeles County, and Baja California.

The remaining 1000 genomes were allocated to all other locations proportionally to their distance to San Diego and the total number of flights connecting the location and San Diego in 2019. Here, location refers to a state (or first administration level) in the US and Mexico, and country everywhere else. Geographic distance was calculated as the centroid-centroid distance to San Diego County, rescaled to have unit scale, and inverted, so that nearby locations had the greatest value. Total number of flights into San Diego was obtained from the OpenSky Network [56], and also rescaled to have unit scale (for more details see following methods section *Travel and Mobility Data)*. The sum of these two values proportional to all other locations was the proportion of the 1000 contextual genomes allocated to that location. In order to sample virus diversity in each location equally, sequences were randomly sampled proportional to the location-specific incidence data binned by epidemiological week.

To estimate a root with a reasonable date and location state in our phylogenetic inference, we also included the 50 earliest SARS-CoV-2 genomes in our dataset. To accurately infer the timing and geographic state of the lineages responsible for widespread epidemiological waves, we included the 10 earliest sequences assigned to Alpha, Delta, BA.1(Omicron) and BA.2 (Omicron). Lastly, to assess the accuracy of the timing of the basal structure of our phylogeny, we included genomes from three outbreaks with well-described introductions[7,14,57]. A list of all included sequences, their GISAID accession IDs, and the compartment they filled is shown in Supplemental Table 1.

### Phylogenetic Analysis

We aligned the sequence dataset to reference genome Hu-1 (GISAID ID: EPI_ISL_402125) using minimap2 v2.17 and gofasta v0.0.6(<u>virus-evolution/gofasta</u>)[58]. We masked the 3’ and 5’ UTRs as well as sites that may confound phylogenetic inference of SARS-CoV-2 genomes[59]. We constructed a maximum likelihood phylogenetic tree for the dataset using IQ-TREE2 and an HKY substitution model[60,61]. We rooted the resulting phylogeny on Hu-1 and time-resolved it using TreeTime v0.7.4 with a strict clock rate of 0.00091 substitutions/site/year, pruning taxa that were more than three interquartile ranges from the clock-rate regression[62]. Lastly, we randomly resolved polytomies in the tree by adding 0 length branches with gotree[63].

We reconstructed the time-resolved phylogeny using BEAST v1.10.15[64]. We used the HKY substitution model with gamma distributed rate variation among all sites. We fixed the clock rate at 9.1x10^-4^ substitutions/site/year and used an exponential growth coalescent tree prior. We also fixed the root of the tree on November 20th, 2019[65]. We combined two independent MCMC chains of 200 millions states ran with the BEAGLE computational library[64]. Parameters and trees were sampled every 10,000 and 100,000 steps, respectively, with 20-60% of steps discarded as burn-in (depending on the chain). Convergence and mixing of the MCMC chains were assessed with Tracer v.1.7.2, and all estimated parameters were determined[66] to have effective sample sizes of greater than 100.

### Phylogeographic Reconstruction

We performed two discrete state ancestral reconstructions on geographic states using BEAST. This analysis reconstructed location-transition history across an empirical distribution of 2000 time-calibrated trees sampled from the posterior tree distribution estimated above. In the first analysis the discrete states used were (1) San Diego County, (2) Los Angeles County, (3) USA (not including either California county), (4) Baja California, (5) Mexico (not including Baja California), and (6) a final state corresponding to all remaining locations. The second analysis assigned San Diego County taxa into the County of San Diego Health and Human Services (HHSA) region they were collected in based on the ZIP code they were collected from. The ZIP code to HHSA region table we used was retrieved from HHSA website (https://www.sandiegocounty.gov/content/dam/sdc/hhsa/programs/sd/community_action_partnership/26%20HHSA%20sdcnty_zipcode.pdf). We assumed that geographic transitions rates were reversible and used a symmetric substitution model for both analyses. We used Bayesian stochastic search variable selection to infer non-zero migration rates[35]. We used the TreeMarkovJumpHistoryAnalyzer from the pre-release version of BEAST v1.10.5 to obtain the Markov jump estimates and their timings from the posterior tree distribution, and assumed that they are a suitable proxy for the transmission between two locations[35,67],[68]. We used TreeAnnotator v1.10 to construct a maximum clade credibility (MCC) tree which we visualized with baltic (https://github.com/evogytis/baltic).

### Persistence Analysis

We used the PersistenceAnalyzer from the pre-release version of BEAST v1.10.5 to summarize the relative contribution of independent introductions on local circulating lineages in San Diego across the posterior tree distribution labeled with Markov jumps. Briefly, for each two week period represented in the phylogeny, we identified the number of lineages circulating in San Diego at the end of the period and determined whether they resulted from a lineage that was estimated to be circulating in San Diego at the beginning of the period or from a unique introduction during the period. Persistent lineages are lineages that could be traced back to locally circulating lineages.

### Contact Tracing Data

Contact tracing data was provided to us by the Epidemiology and Immunization Service Branch of the County of San Diego. Up until March 2022, contact tracers in San Diego interviewed between 40-60% of all confirmed cases in the county and asked, among other questions, whether there was travel within the US (excluding California), Mexico, or internationally during the 2-16 days prior to onset of symptoms (or positive test date if asymptomatic). Uncertainty in the proportion of interviewed cases that were travel-related was assessed by bootstrapping interviews for each week 100 times.

### Travel and Mobility Data

We followed Zeller et al.[14] in calculating travel into San Diego County, using the weekly patterns data from SafeGraph, a data company that aggregates anonymized data from numerous applications to provide insights about physical places, via the Placekey Community (See Zeller at el. citation for full calculation details). SafeGraph estimates human movements using cell phone tracking, which has been shown to capture both land and air travel at a variety of distance scales[2,69]. Briefly, we estimated the true number of travelers for a given week (*w*) between a source and destination location (*S* and *D*; travelers_w,S>D_) using the raw number of devices that traveled from the source to the destination location (*devices_w,S>D_*), the total number of devices detected at the destination(*totalDevices_w,D_*) and the total population of the destination location (*population_D_*), according to:

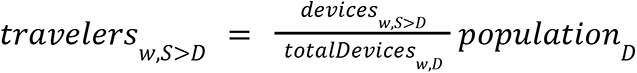

We note that because of the EU General Data Protection Regulation, SafeGraph was not able to provide mobility data from countries within the European Economic Area (see https://www.safegraph.com/privacy-policy). Therefore, EU countries were missing from our mobility analyses. We also note that SafeGraph does not provide mobility data finer than the country-level for international locations, particularly Mexico. However, independent travel surveys indicate that it is reasonable to assume that 99% of travel into San Diego from Mexico originated in Baja California[36].

We noticed that travel from international locations from 2020 onwards increased uniformly relative to data from 2019. This was not consistent with independent sources of mobility. For example, we observed no increase in monthly inbound crossings at the US-Mexico border into San Diego collected by the Department of Transportation (https://explore.dot.gov/#/views/BorderCrossingData/Monthly). To correct this artifact, we normalized mobility data from January 1^st^, 2020 onward by multiplying it by the ratio of the mean mobility between January 1^st^ and March 1^st^ in 2019 compared to 2020. March 1^st^ was chosen because it was generally before any reductions in mobility occurred as a result of the spread of SARS-CoV-2 in the US. The impact of this correction should be slight as our conclusions rely on the relative, rather than absolute mobility, into San Diego.

We also obtained weekly air travel flight data into San Diego International Airport (KSAN) from the OpenSky Network[56]. We filtered data for flights with complete origin and destination airport ICAO codes. ICAO codes were matched to country, state, and county location using an airport database (https://github.com/mwgg/Airports). Using the dataset, we counted the total flight counts from each US state and non-US country in 2019 to use as an input for the genomic database generation.

### Epidemiological Data and Estimated import Risk

We calculate import risk according to du Plessis et al.[15], with some modifications. Briefly, we estimated the number of infected travelers arriving each day into San Diego from each source location as the product of the number of asymptomatically infectious individuals in each source location on that day and the number of travelers arriving in San Diego from the source location as estimated from the SafeGraph mobility data. SafeGraph mobility data utilizes cell phone tracking data so both air and land travel is included. Similar to du Plessis et al., we conservatively estimate that only asymptomatic infections contributed to imporation risk, as symptomatic infections would not travel. Therefore, the asymptomatic infection rate is derived only from the number of pre-symptomatic and asymptomatic infections.

We estimated the asymptomatic infectious rate for each location by back-extrapolating the death time series assuming the same estimates for the latent and incubation period, infectious duration, symptom-onset-to-death and asymptomatic proportion as du Plessis et al.[15] (See citation for full details). We also note that infectious rates were back-calculated from reported deaths using a constant infection fatality rate, even though it likely varied between locations and across time[70]. For instance, death ascertainment rates were known to be lower in Baja California[71] than in California[72]. Conclusions were primarily drawn from temporal dynamics rather than absolute values so the effect of this bias should be minimal, but results drawn from the latter should be viewed with some uncertainty.

Because travel surveys indicated that 99% of all Mexican travelers visiting San Diego originated in Baja California, we used Baja California’s asymptomatically infectious individuals in place of Mexico’s for estimating import risk[36]. We obtained the time series of reported deaths from each California county, US state, and county from the outbreak.info R package which provides data from the COVID-19 Data Repository by the Center for Systems Science and Engineering (CSSE) at Johns Hopkins University[73,74]. We additionally obtained the time series of reported deaths from each Mexican state directly from the Mexican Department of Health (https://datos.covid-19.conacyt.mx/), as they were more complete than other sources.

Counterfactual import risk was calculated as above, except that for dates from March 1^st^, 2020 onward the number of travelers arriving in San Diego on a given day were replaced with the number of travelers arriving in San Diego for that day in 2019.

### Code and Data Availability

Code for all analyses and figure generation, XMLs and log file for BEAST analyses, and configs for simulations are available at: https://github.com/andersen-lab/project_2023_SARS-CoV-2_Connectivity. Sequencing data, including consensus sequences and raw data, is available on NCBI under the BioProject accession ID PRJNA612578. Raw sequencing data is also available on our <u>Google Cloud</u>.

## Data Availability

The data that support the findings of this study are openly available in Github at https://github.com/andersen-lab/project_2023_SARS-CoV-2_Connectivity

## Acknowledgements

We thank the administrators of the GISAID database for supporting rapid and transparent sharing of genomic data during the COVID-19 pandemic and all our colleagues sharing data on GISAID. The research leading to these results has received funding from the National Institutes of Health (grants U19AI135995, U01AI151812, UL1TR002550, F32AI154824, and R01AI153044) and the CDC contract 7530122C14843. We also gratefully acknowledge support from NVIDIA Corporation and Advanced Micro Devices, Inc., with the donation of parallel computing resources used for this research.

## Author Contributions

Conceptualization, GWH, KG, JIL, EP, XJ, MW, KGA, SW, MZ; Methodology, NLM, GWH, KG, SW, MZ; Software, GWH, KG; Formal Analysis, NLM, GWH; Investigation, NLM; Resources, EK, MAS, SAP, DP, AH, PD, WC, AC, AR, AV, AR, CW, JH, JW, SNN, AP, ESL, EWS, ERE, RT, MC, NAB, PS, RAS, SA, TTN, TB, TO, RF, EHS, OEZ, IS, MS, JLM, AG, ARM, EM, JC, JDM, SS, SS, NM, AW, CA, ECH, JJL, KSR, KNN, KS, KMF, SLG, AB, DM, SK, NKM, RTS, AJN, HJR, JRC, MLG, JMC, DB, MI, NLW, WL, RSG, MAL, JA, BH, KJ, BO, GB, IL, JER, RF, SFK, AS, AP, BW, GM, MH, RWM, SLR, TW, IHM, RAF, LN, MMQ, AH, AM, OB, SHB, JO, DGR, MG, LCR, BJA, JG, AH, RK, LCL, GWY, AC, MZ; Data Curation, NLM, EK, MAS, SAP, OEZ, IS, MS, JLM, AG, NM, AW, CA, CMA, ECH, JJL, KSR, KNN, KS, KMF, AB, AJN, HJR, JRC, MLG, JMC, DB, MI, NLW, WL, MAL, JA, GB, JER, RAF, JO, DGR, MG, JG, AC, MZ; Writing - Original Draft, NLM; Writing - Review & Editing, NLM, KG, JIL, EP, KGA, SW, MZ; Visualization, NLM; Project Administration, EGS, RR; Funding Acquisition, KGA; All authors contributed to interpreting and reviewing the manuscript.

## Declaration of Interests

K.G.A. has received consulting fees and compensated expert testimony on SARS-CoV-2 and the COVID-19 pandemic.

## Supplemental Figures

**Supplemental Figure 1.**
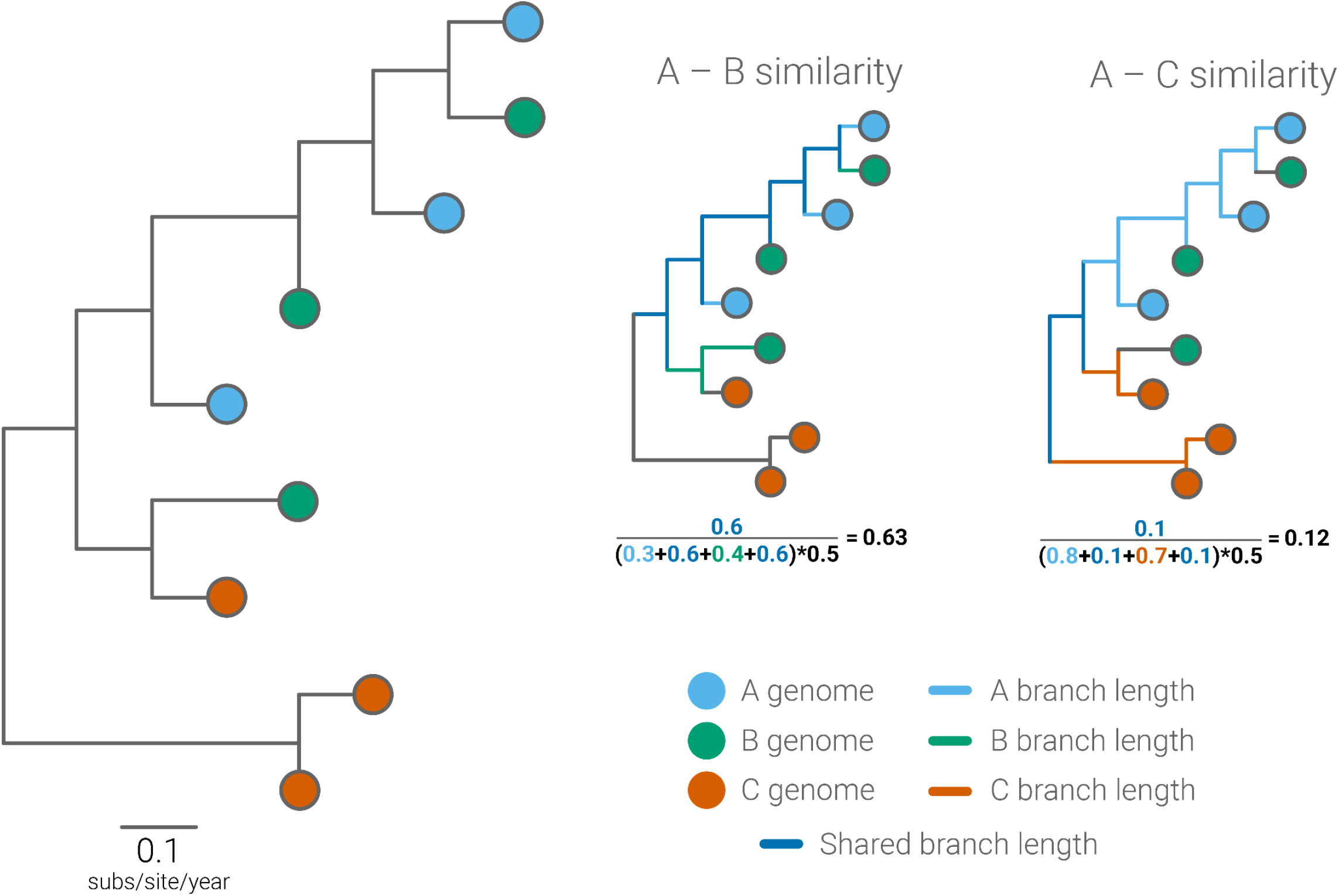
Diagram of the PhyloSor metric. PhyloSor quantifies the phylogenetic similarity of two communities as the proportion of branch lengths that are shared by the communities compared to the total branch lengths of both communities. PhyloSor similarity ranges from 0, where two communities share only a very small root, to 1, where two communities have identical taxa. In this example tree, scaled by substitutions/site/year, communities A and B are more similar than communities A and C as indicated by the higher PhyloSor value. Values in the PhyloSor formula are colored by the branches they summarize.

**Supplemental Figure 2.**
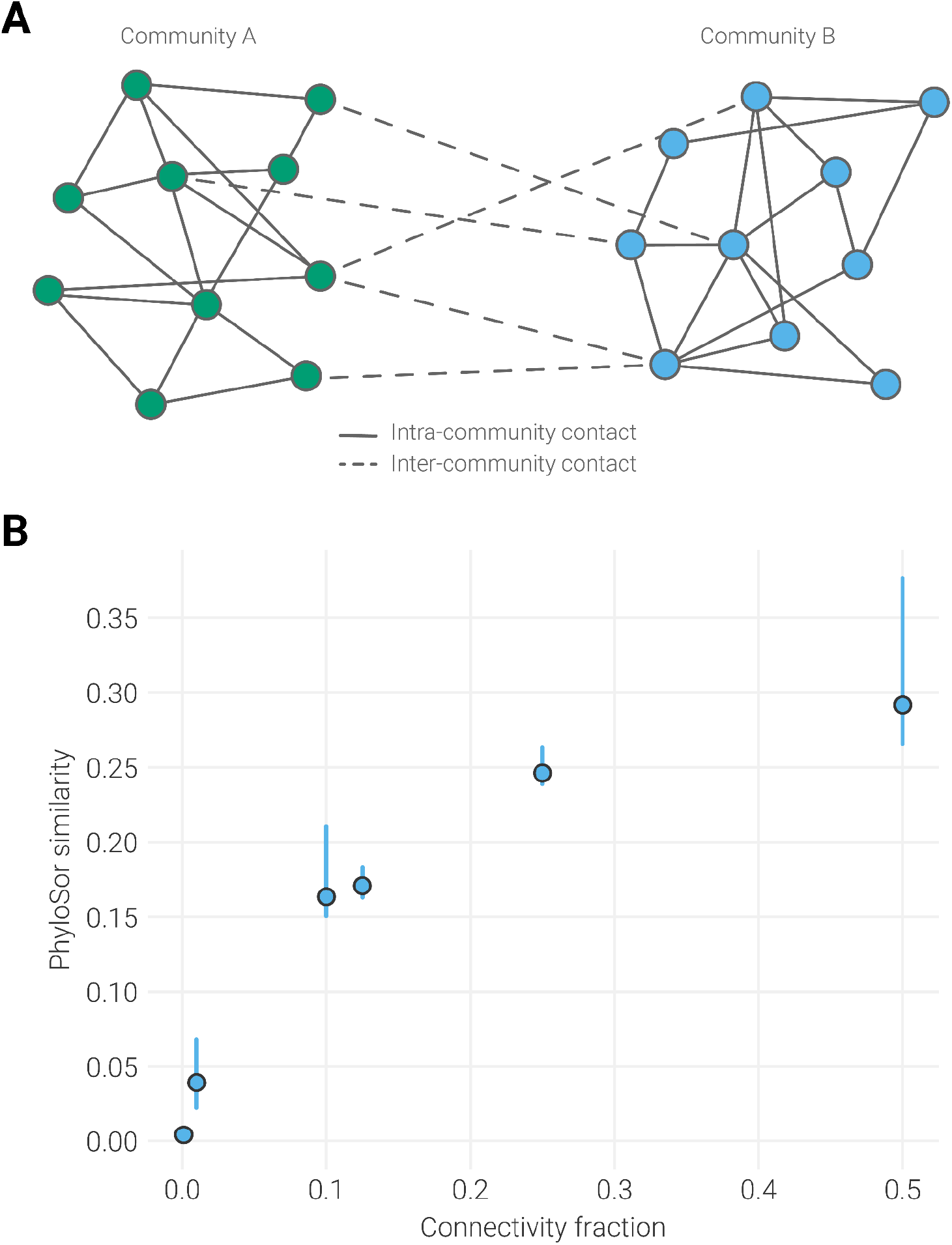
PhyloSor recapitulates connectivity between communities in simulated phylogenies. (**A**) Example bipartite contact network containing communities with size 10. Nodes are colored based on their membership in one of two communities. Connectivity fraction is calculated as the ratio of inter-community contacts (dashed edges) to the average intra-community contacts (solid edges). Shown connectivity fraction is 5 (inter-community edge) / 17 (average inter-community edges) = 0.3. **(B**) PhyloSor similarity between two communities with the indicated connectivity, which represents the ratio of inter-community edges to mean intra-community edges in the simulated contact network. To limit temporal variation, only PhyloSor similarity from the first month of the simulated phylogeny is shown. Distributions represent the range of values from 10 independent simulations. A strong significant correlation was found between connectivity fraction and PhyloSor similarity (Pearson r = 0.89 [95% CI: 0.82-0.93]; P < 0.001).

**Supplemental Figure 3.**
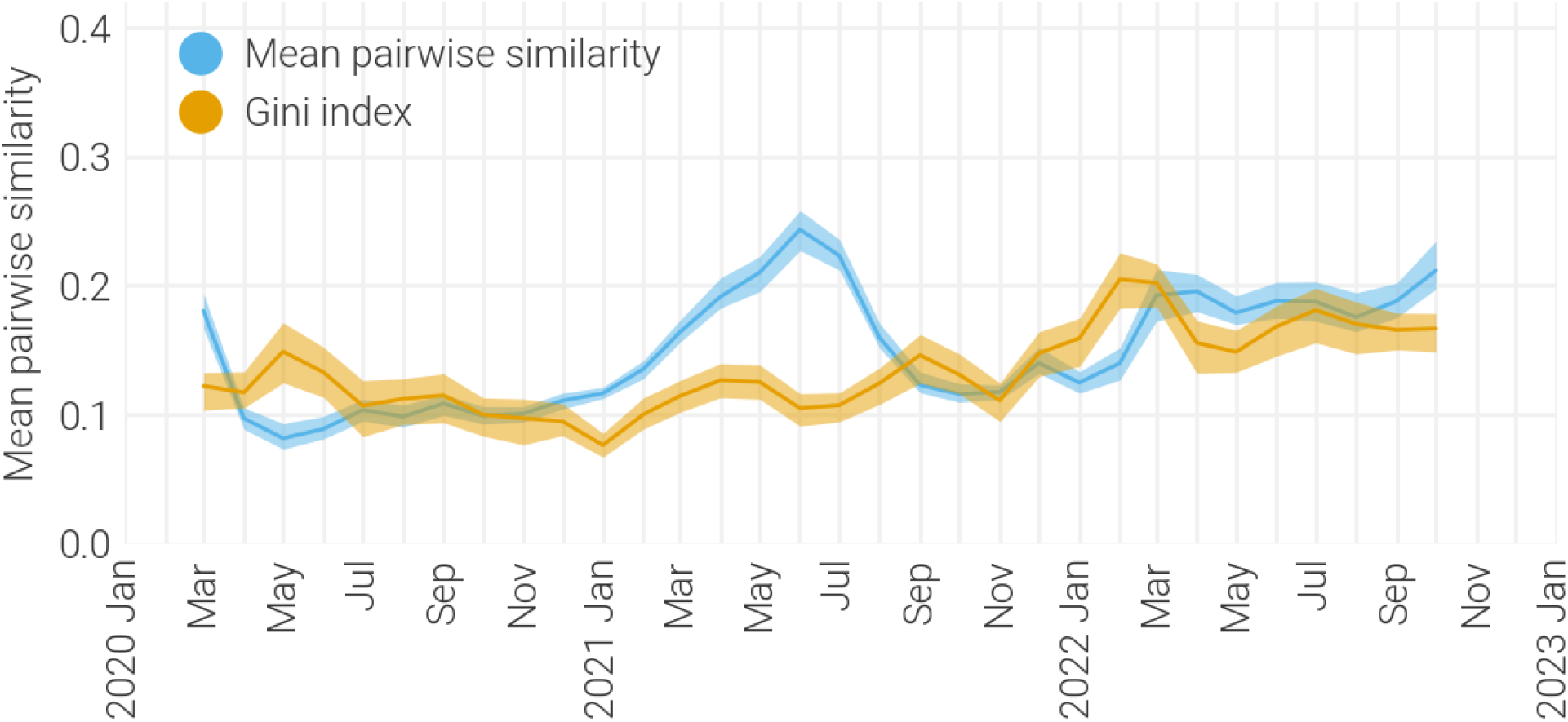
Heterogeneity in North American locations’ contribution to graph efficiency. Temporal trends in the contribution of each location to graph efficiency as measured by Gini index (indicated by orange line). 95% confidence intervals were calculated for each month by bootstrapping nodes in the network 100 times. Temporal trends in graph efficiency are indicated by the blue line and represent the same data as in **Figure 1A**.

**Supplemental Figure 4.**
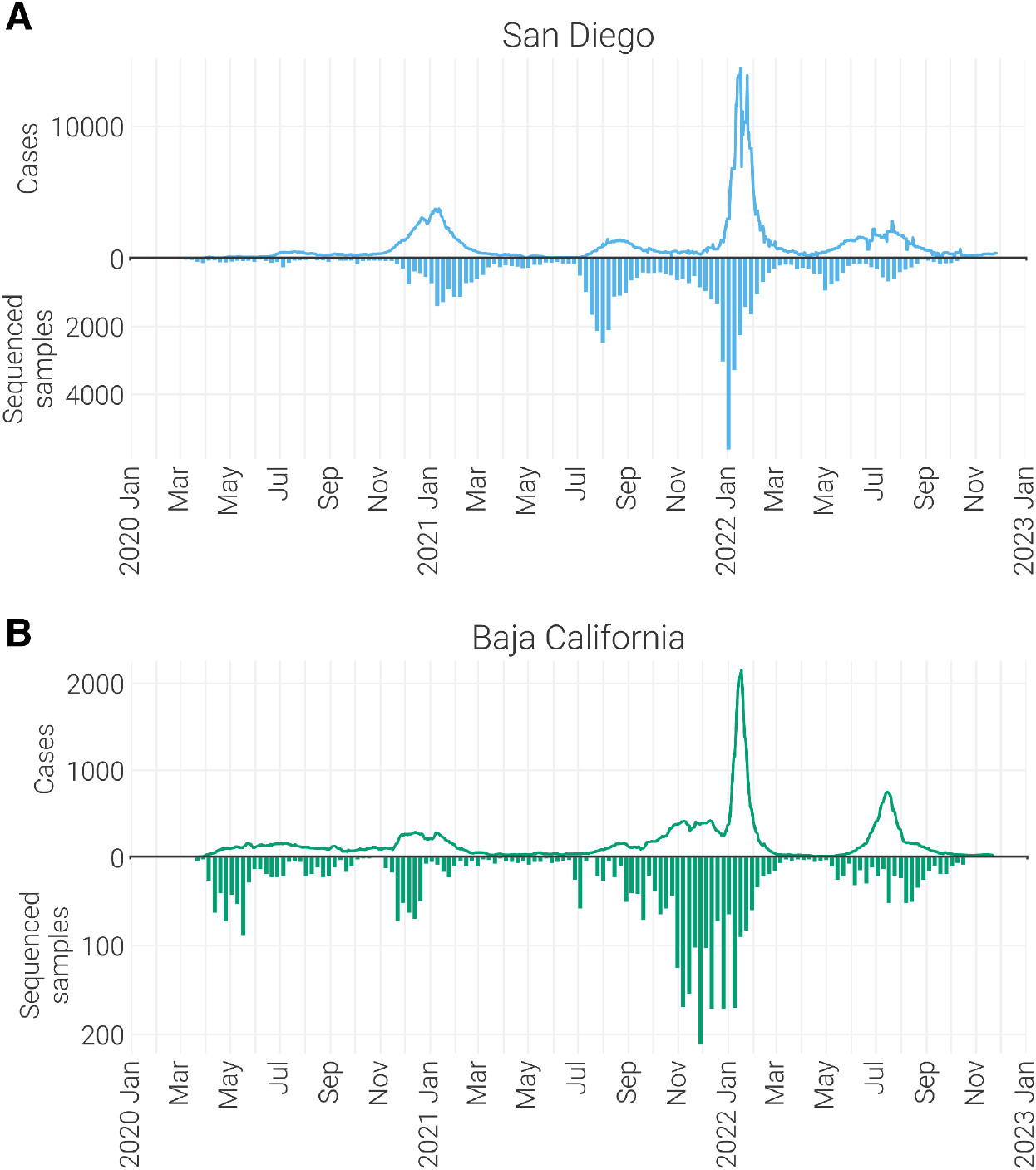
Genomic sampling in San Diego and Baja California. (**A**) Top axis indicates the 7-day rolling average of daily reported cases in San Diego, while the bottom axis indicates the number of samples sequenced for each week of the pandemic. (**B**) Same as in A, but for Baja California.

**Supplemental Figure 5.**
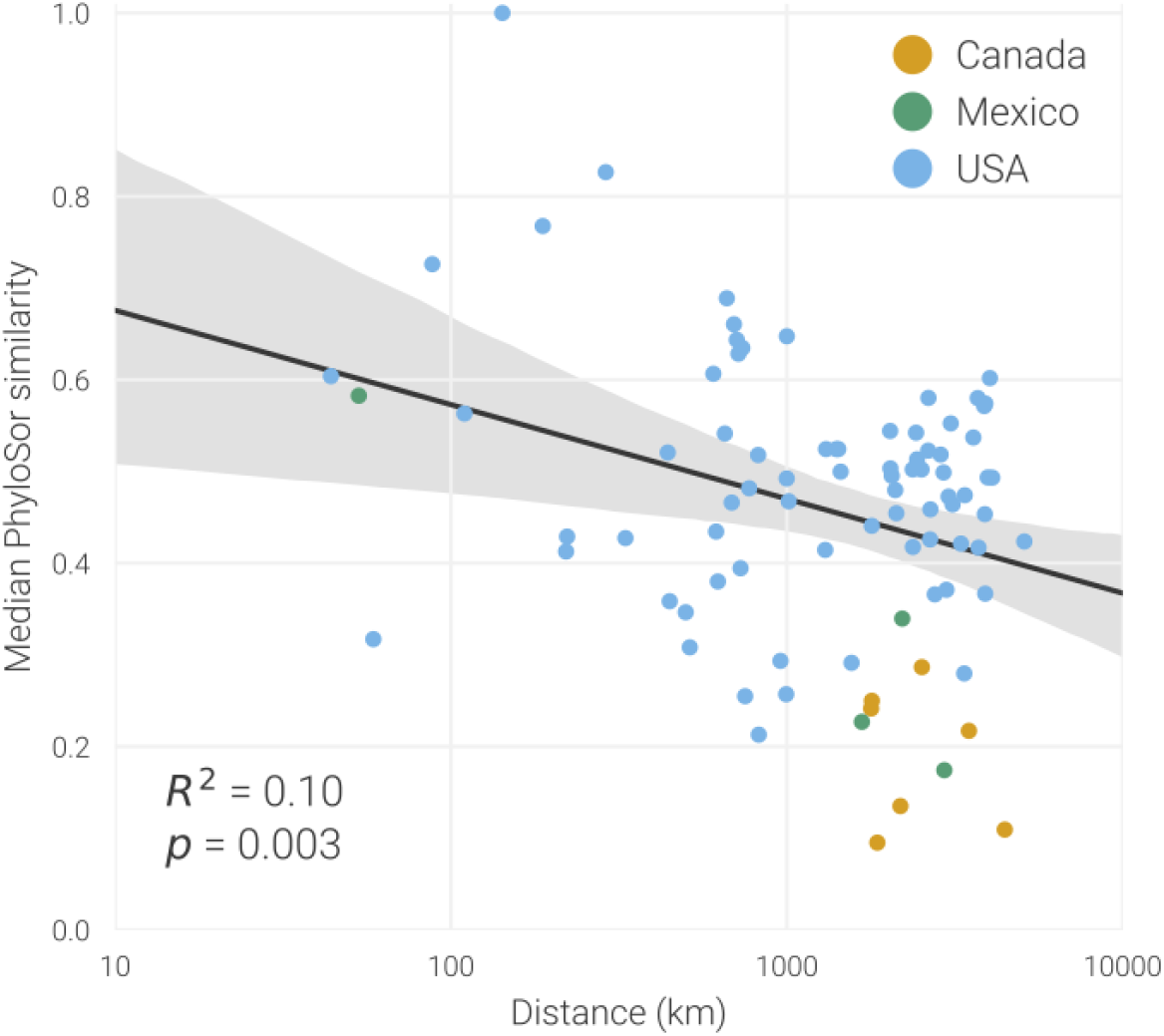
PhyloSor similarity is weakly explained by geographic proximity. Relationship between each locations’ median normalized PhyloSor similarity to San Diego and their log-transformed centroid-centroid distance to San Diego. Locations are colored based on their country (Canada - orange, Mexico - green, USA - blue). Strength of correlation was determined using Spearman’s correlation coefficient.

**Supplemental Figure 6.**
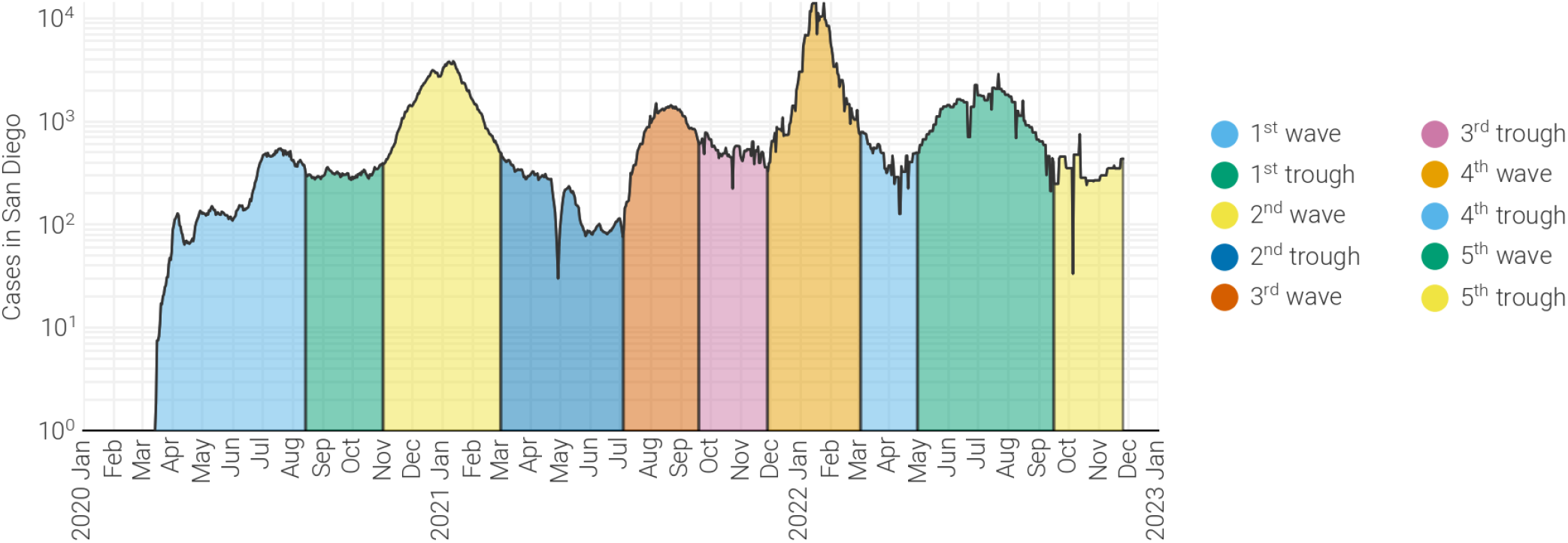
Epidemiological waves in San Diego. Daily reported cases in San Diego are annotated according to their epidemiological phase.

**Supplemental Figure 7.**
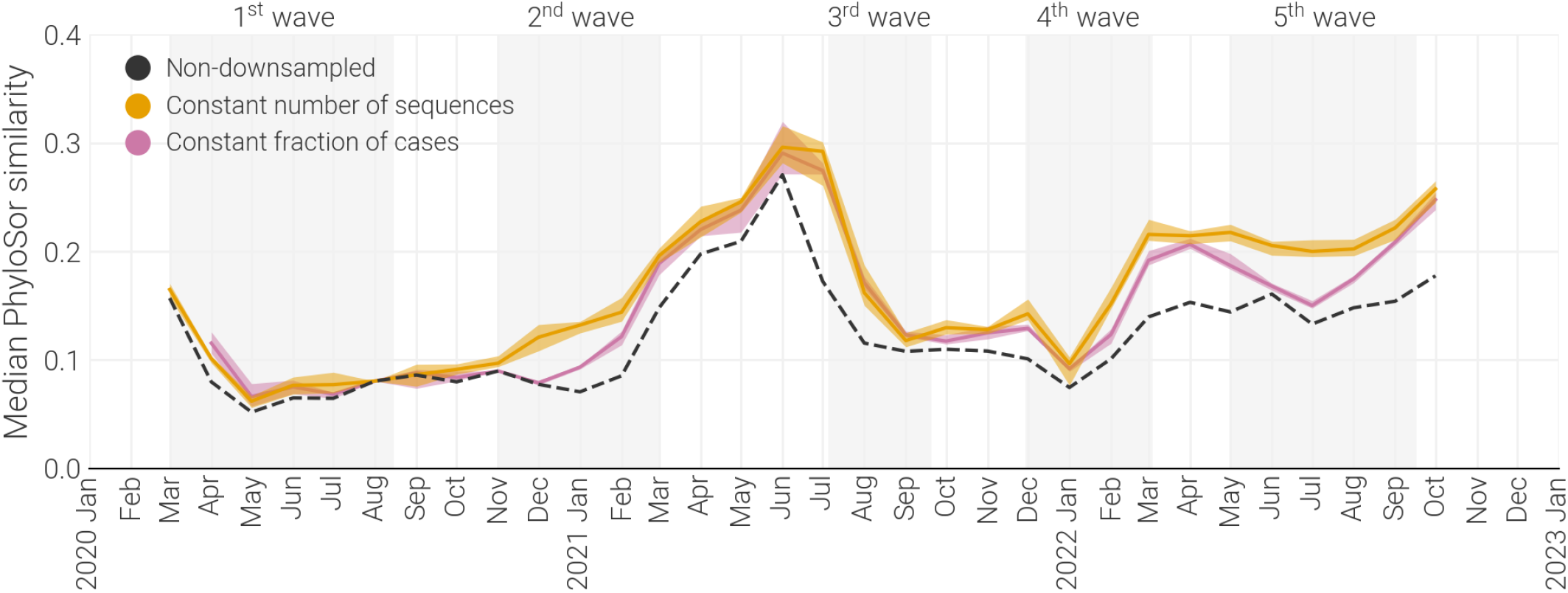
Trends in PhyloSor similarity are independent of sampling. San Diego’s median PhyloSor similarity to all other location when all sequences are included (black dashed line), a constant number of San Diego sequences are included from each month (orange line), or constant proportion of sequences relative to cases are included from each month (magenta line). Subsampled results display 95% confidence intervals calculated from 10 independent subsamplings. Subsampled results are strongly correlated with and display low variance in differences to the non-downsampled results (minimum Spearman r^2^ = 90.5% for a constant number of sequences and 93.1% for a constant fraction of cases; Root-mean-square error to non-downsampled results = 0.048 for a constant number of sequences and 0.036 for a constant fraction of cases).

**Supplemental Figure 8.**
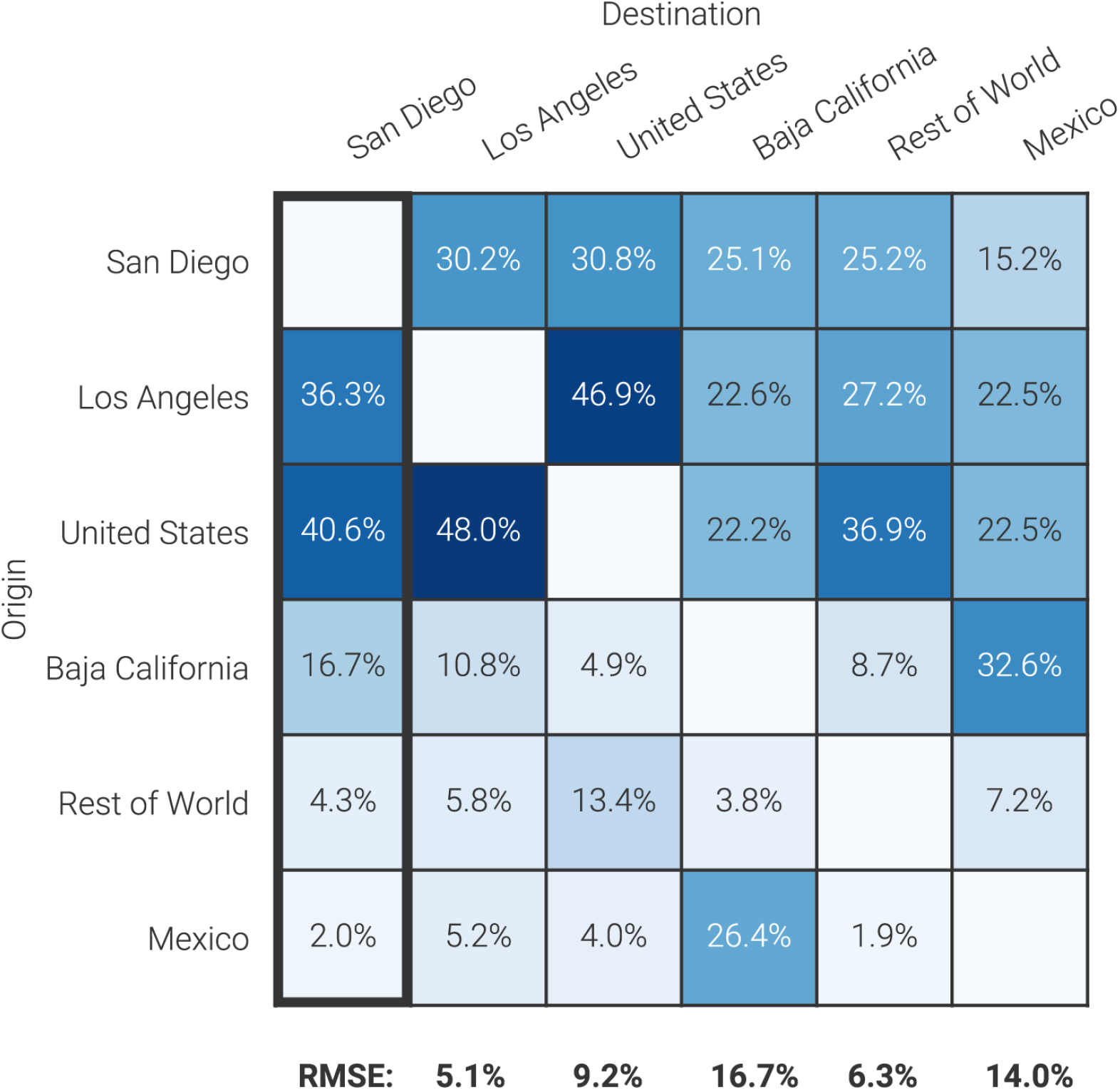
Schematic of the comparison of introductions profiles. Example data showing the proportion of introductions into each location state that originated in each other state (i.e. an introduction profile). Columns may not sum to 100% due to rounding. Each locations’ introduction profile was then compared to San Diego’s using Root-mean-square error (indicated below each column).

**Supplemental Figure 9.**
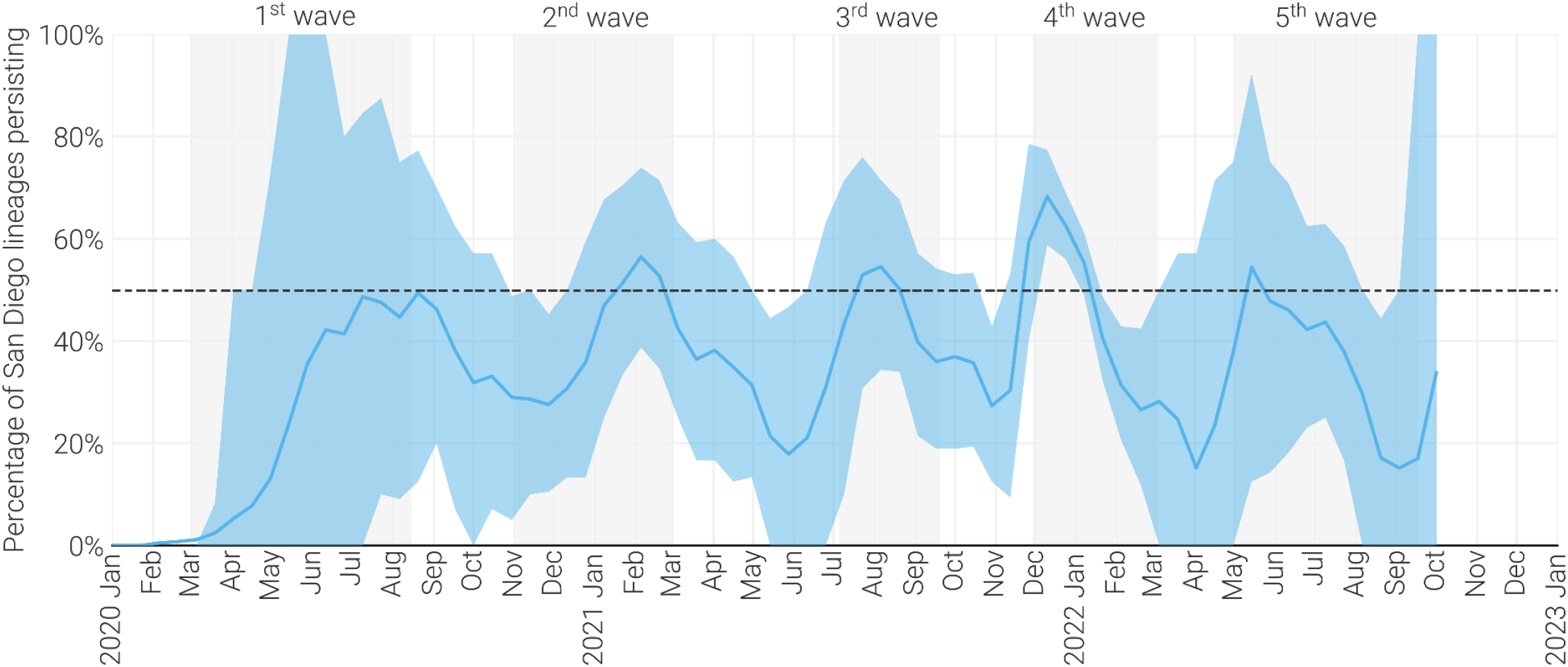
Persistence of San Diego lineages over time. Percentage of unique San Diego lineages that are estimated to have persisted in San Diego since two weeks prior, for each non-overlapping two week period between January 2020 and October 2022. Dashed line indicates persistence of 50% of lineages.

**Supplemental Figure 10.**
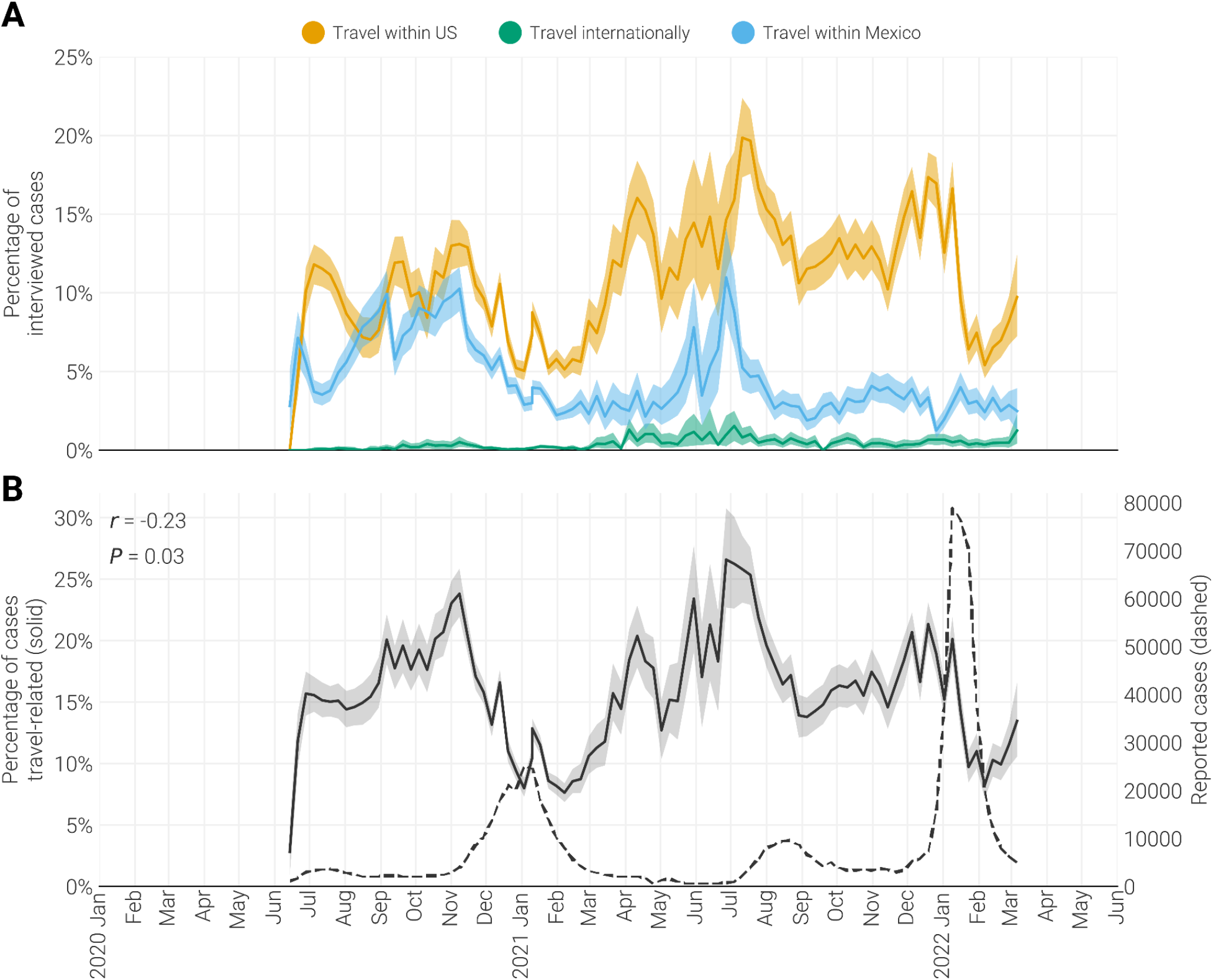
Travel associated cases in San Diego. (**A**) Percentage of interviewed cases that reported travel within the US, Mexico, or internationally, during the 2-14 days prior to their onset of symptoms (or specimen collection if asymptomatic). Confidence intervals were calculated by bootstrapping interviews 1000 times. (**B**) Percentage of interviewed cases that reported any travel (solid line) compared to the weekly number of reported cases in San Diego (dashed line). A significant negative correlation was found between the reported number of cases and the percentage of cases that were travel related (Pearson r = -0.23 [95% CI: -0.03 to -0.42]; P = 0.03).

**Supplemental Figure 11.**
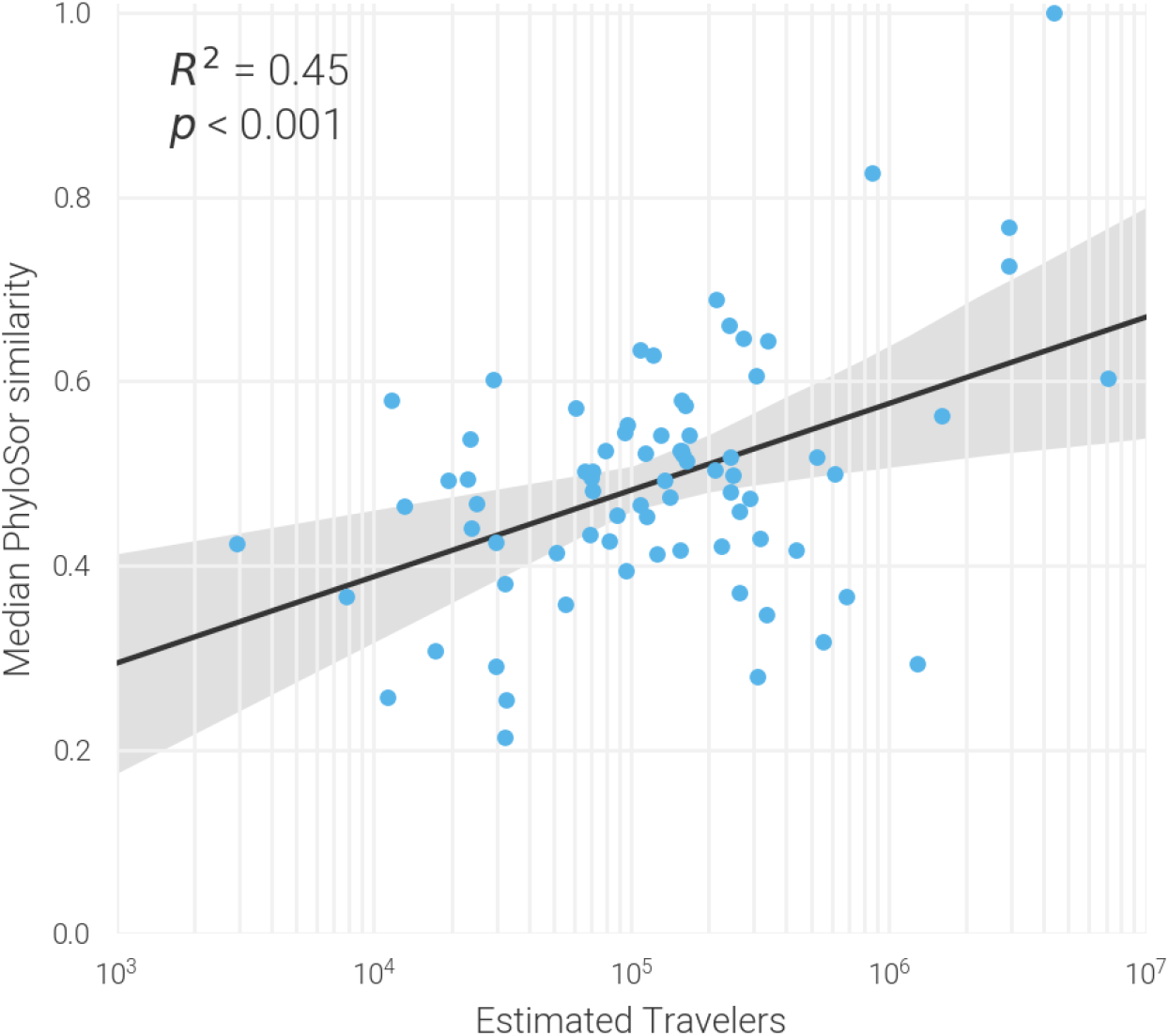
Phylosor similarity is explained by mobility. Relationship between each locations’ median normalized PhyloSor similarity to San Diego and their log-transformed total number of total travelers to San Diego from January 2020–June 2021. Strength of correlation was determined using Pearson correlation coefficient.

**Supplemental Figure 12.**
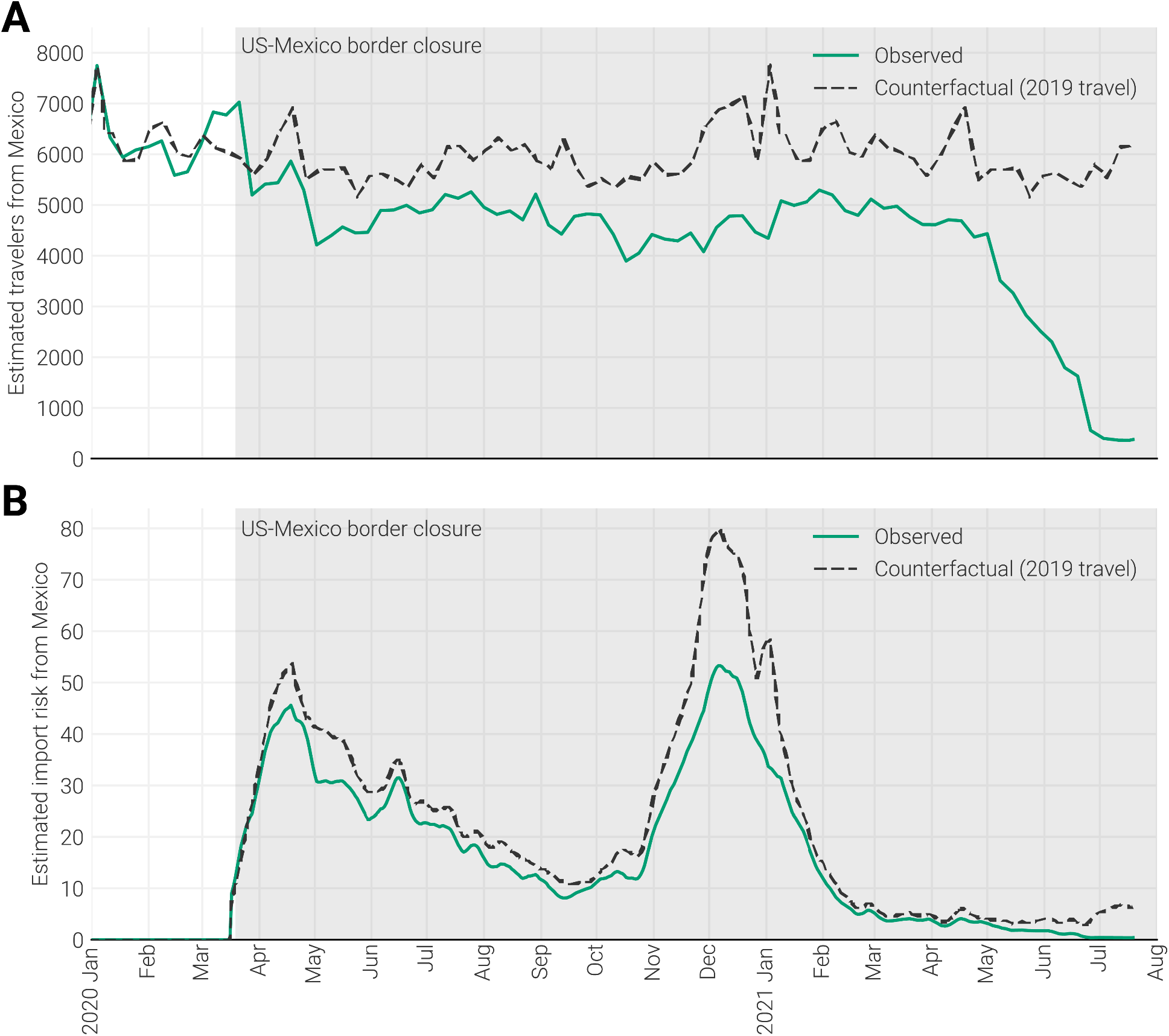
The effect of the border closure on import risk from Mexico. **(A)** Estimated number of travelers into San Diego from Mexico. Green curve indicates observed travel volume, and dashed black curve indicates counterfactual where travel volumes from 2019 was extended to 2020-onwards. **(B)** Import risk into San Diego from Mexico estimated using observed travel data (green curve) or assuming travel volume from 2019 was extended to 2020-onwards (counterfactual; black dashed curve).

## Notes

### Author Declarations

Sample collection, RNA extraction, and viral sequencing was evaluated by the Institutional Review Board (IRB) at Scripps Health (IRB-21-7739). All samples were de-identified before receipt by the study investigators. Aggregated contact tracing data was publicly available prior to the initiation of the study.

